# Quantifying advances in infectious disease forecast skill across seasons and pathogens

**DOI:** 10.64898/2026.09.29.26364307

**Authors:** Clara Bay, Nima R. Moghaddas, Jessica T. Davis, Rebecca K. Borchering, Matthew Biggerstaff, Alessandro Vespignani, Michael A. Johansson

## Abstract

Infectious disease forecasting supports public health preparedness and response by informing resource allocation, intervention planning, and situational awareness. Infectious disease forecasting capabilities in the United States have grown over the past decade yet whether forecast quality has improved over this time remains unclear. Current evaluation metrics describe relative performance for a common set of forecast targets but cannot evaluate performance across different outcomes, seasons, and pathogens. We addressed this challenge by developing a novel skill score metric to assess forecasts based on performance relative to two benchmark models: a hindcast as an idealized upper bound of performance and a baseline model as a minimal performance standard. To study the impact of the baseline choice, we considered three types of baseline models with varying levels of complexity. Using this skill score framework, we retrospectively evaluated short-term influenza (influenza-like illness and hospitalization) and COVID-19 (case, death, and hospitalization) forecasts, focusing on how the performance of team and multi-model ensemble forecasts changed over time. The ensemble outperformed 99% of forecasts from a simplistic, naïve baseline, and 84-91% of forecasts from more complex baseline models that incorporate recent data. We found consistent improvements in ensemble and team forecast skill over time for both influenza forecast targets, with 4-week horizon forecasts in the most recent year having higher skill scores than 1-week horizon forecasts in the first year of each challenge. Results were mixed for COVID-19 forecasts. This framework provides a comprehensive picture of progress in infectious disease forecasting over the past decade enabled by improved standardization, data, models, and collaboration.

## Introduction

Over the past decade, infectious disease forecasting has developed into a formalized, collaborative, operational endeavor. Forecasts have been generated in response for seasonal influenza, COVID-19, dengue, Ebola, and West Nile virus [16, 24, 30, 37, 58]. The ability to predict the trajectory of an ongoing outbreak is central to public health preparedness and response, yet the usefulness of a forecast depends on the quality of its predictions. Reliable predictions of outcome targets such as cases, deaths, or hospitalizations allow policymakers and healthcare workers to anticipate surges, allocate resources, plan timely interventions, and communicate risk to the public [5, 15, 47].

In the United States, coordinated routine and real-time forecasting began in 2014 when the Centers for Disease Control and Prevention (CDC) launched the Epidemic Prediction Initiative, leading to the start of the FluSight Forecasting Challenge during the 2014-15 influenza season, with the goal of forecasting seasonal influenza dynamics [3, 4, 10, 23, 37]. The COVID-19 pandemic prompted rapid expansion of this infrastructure, with the CDC-supported COVID-19 Forecast Hub collecting and analyzing case, hospitalization, and death forecasts from over 50 modeling teams beginning in April 2020 [5, 17]. These initiatives have produced forecasts spanning multiple pathogens, seasons, and surveillance systems, and have driven innovation and adoption of infectious disease forecasts by public health agencies. Among key advances were the implementation of probabilistic forecasts in common forecasting formats, standardized evaluation metrics, and the development of baseline models. In addition to facilitating model comparison, these led to the development and evaluation of ensemble forecasts, which have proven to reliably outperform baseline forecasts and the majority of team submitted forecasts [16, 23, 35, 57]. The relatively strong performance of ensembles has led to their use by public health agencies and integration into public facing dashboards.

However, key questions remain. Proper scores, such as the weighted interval score and the logarithmic score, incentivize honest probabilistic forecasting because an optimal score is only mathematically possible if the true observed outcome is predicted [7, 20]. While proper scores reliably differentiate performance by different models on the same outcomes, they are generally not useful as absolute performance measures across different forecasted outcomes or to classify forecasts as good or bad. This limits the ability of proper scores to help us understand if forecast models are getting better as infectious disease forecasting has evolved and how much better forecasts could get. For example, a season with an atypically high peak or noisier data will generally yield worse proper scores compared to forecast scores for the same model in a more typical season. Assessing performance across these contexts risks confounding forecast model quality with underlying outcome variability and requires a framework that measures performance relative to benchmarks that account for these factors.

To address these challenges, we assessed performance by developing a novel skill score that analyzes performance relative to benchmark models, based on methods used in weather forecasting [42, 60]. Here, we compare forecasts to two types of benchmark models: a hindcast, which represents an idealized ceiling of forecast performance accounting for noise in the observed data, and a baseline model, which serves as a minimal performance standard, using 3 different baseline models as lower performance bounds. These benchmarks improve the interpretation of forecast performance and serve as upper and lower bounds of expected forecast quality.

With this novel skill score framework, we retrospectively evaluated over 2.5 million influenza and COVID-19 forecasts in the United States to characterize forecast performance over time across these efforts. We examined how performance varies across forecast horizon, how the choice of reference models and scoring metrics affect evaluation, and how forecast skill has evolved across influenza and COVID-19 forecasting challenges. These analyses provide a comprehensive picture of the current state of infectious disease forecasting.

## Results

### A decade of forecasts

We evaluated forecast performance for two influenza targets (weekly % outpatient influenza-like illness visits (ILI) from 2015-16 – 2019-20 and weekly hospitalizations from 2021-22 – 2025-26) and four COVID-19 targets (weekly cases in 2020–2021, weekly deaths in 2020– 2023, daily hospitalizations 2021–2023, and weekly hospitalizations in 2025), described in Table 1. These six targets capture different aspects of respiratory disease burden, endemic and emergence dynamics, include 2,527,077 publicly available forecasts over 19 target-year-season combinations, and cover ten calendar years of infectious disease forecasting efforts. Most forecasts were generated weekly at 1-through 4-week-ahead horizons relative to the most recently available outcome data at the time the forecast was made. While recent COVID-19 and influenza hospitalization forecast challenges have defined the horizon relative to the forecast date, we adopt the earlier definition of the horizon relative to available data for comparison across challenges. COVID-19 hospitalization forecasts used daily data and daily forecast horizons from 2021-2023; we analyzed the Saturday outcomes, aligning the forecast horizons with the other challenges, and excluded 2024 COVID-19 hospitalizations forecasts, when the data and target switched from daily to weekly and there was a 6-month data availability gap. We analyzed influenza forecasts by season, typically October-May as defined in the challenges, and COVID-19 forecasts by calendar year as forecasts were produced year-round.

**Table 1:** Infectious disease forecasts included in the analyses.

| Disease | Target | Seasons | Geographic Level | Submission Format | Forecasts Evaluated |
| --- | --- | --- | --- | --- | --- |
| Influenza [13] | Weekly % of outpatient ILI visits | 2015–16 – 2019–20 | U.S. HHS regions & national | Binned probability distributions (2015–16: 0.5% bins, 2016–17 – 2019–20: 0.1% bins) | 145,676 |
| Influenza [9, 12] | Weekly incident hospitalizations | 2021–22 – 2025–26 | U.S. states, District of Columbia, Puerto Rico, & national | 23 quantiles (1, 2.5, 5, 10, 15, ..., 85, 90, 95, 97.5, 99%) | 706,518 |
| COVID-19 [49] | Weekly incident cases | 2020 – 2021 | U.S. states, District of Columbia, Puerto Rico, Virgin Islands, & national | 7 quantiles (2.5, 10, 25, 50, 75, 90, 97.5%) | 348,952 |
| COVID-19 [49] | Weekly incident deaths | 2020 – 2022 | U.S. states, District of Columbia, Puerto Rico, Virgin Islands, & national | 23 quantiles | 754,744 |
| COVID-19 [49] | Daily incident hospitalizations | 2021 – 2023 | U.S. states, District of Columbia, Puerto Rico, Virgin Islands, & national | 23 quantiles | 496,144 |
| COVID-19 [11] | Weekly incident hospitalizations | 2025 | U.S. states, District of Columbia, Puerto Rico, & national | 23 quantiles | 75,043 |

To compare forecasts, we first standardized the proper score metric across all submitted and ensemble forecasts. ILI forecasts were originally collected as binned forecast probabilities using two different bin sizes and evaluated using the logarithmic score, while COVID-19 and recent influenza hospitalization forecasts were collected as quantile sets (7 quantiles for COVID-19 cases and 23 for all others) and evaluated using the WIS (Eq. 2). In sensitivity analyses, we found that using the same proper score metric and similar forecast formats led to more consistent evaluation results (Fig. S1). We therefore interpolated 23 quantiles from the binned ILI forecasts to score the ILI forecasts using WIS.

### Skill scores

To assess the performance of forecasts across outcomes and seasons, we implemented skill scores which characterize forecast skill relative to benchmark forecasts:

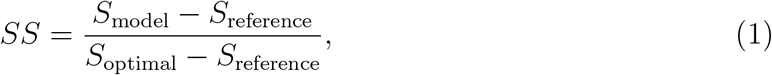

where each *S* is a scoring metric (WIS, here) for a forecast model [60]. The average score of each individual model (*S*_model_) is compared to a reference forecast (*S*_reference_) and to an optimal forecast (*S*_optimal_), all on the same set of forecasting tasks. A score of 1 indicates forecasting equivalent to the optimal model, a score of 0 indicates no improvement from the reference model, and a negative value represents performance worse than the reference model [41, 20]. While skill scores have routinely been employed in weather forecasting and other fields, their use in infectious disease forecasting remains limited. A pairwise relative WIS metric, related to a skill score using perfection as the optimal forecast (WIS = 0) and a challenge-generated baseline forecast as the reference forecast, has been used to show that ensemble and individual team forecasts for COVID-19 and influenza hospitalizations generally outperform baseline models from those challenges [16, 37].

### Benchmark forecasts

The selection of optimal and reference forecasts is critical to skill score interpretation and utility. For the optimal model, one option is to assume that the observed data are the precise true values and that the optimal model predicts them perfectly with zero error and complete confidence [41, 59], as in the pairwise relative WIS [16]. Since epidemiological surveillance systems are intrinsically noisy, incomplete, and variable, we model observation noise using a retrospective hindcast as the optimal model, similar to reanalysis approaches for climate data [51]. Conceptually, the hindcast is a probabilistic estimate of the expected value of the observed outcome given the full data that eventually became available and therefore is not a true forecast and does not vary by horizon. Specifically, we assumed that reported cases, hospitalizations, and deaths followed negative binomial distributions (or logit-normal for ILI) with a mean as a 3-week weighted average of the observed values, centered around the relevant forecast date and the dispersion calculated from the corresponding variance.

Simple, baseline forecasts have been a staple of infectious disease forecasting since structured collaborative forecasting challenges began [4, 44]. We considered three baseline models as standardized skill score reference forecasts: a naïve forecast, a random walk forecast, and a trend forecast. Although these models incorporate varying levels of complexity, they are all simple models that a good forecasting model should be able to outperform.

The *naïve* baseline predicts that future observations will come from the full distribution of past observations, without conditioning on recent data. This is similar to weather predictions based on long-term climate except that here we do not incorporate seasonality as it is not always apparent (e.g., for COVID-19 emergence) and historical data needed to estimate seasonal dynamics are not always available [42]. For each forecast date, we fitted a negative binomial or logit-normal distribution (ILI) to all past data points and used this parametric distribution to generate quantile forecasts for all future observations, regardless of horizon. This represents a simplistic baseline model that we do not expect to generate good predictions when case numbers are high or atypically low. The *random walk* (RW) baseline generates projections anchored at the most recently observed data point, with expanding uncertainty sampled from the one time step differences between all previous observations on the log scale (cases, hospitalizations, or deaths) or logit scale (ILI). The RW baseline is a log-transformed version of the baseline model generated by the COVID-19 Forecast Hub and the FluSight Forecast Hub since 2021, which has an equal probability of increasing or decreasing, resulting in a flat median forecast [44]. The *trend* baseline generates forecasts that follow the trajectory of recent data. We use an exponential smoothing model with additive errors and a damped additive trend [28]. Both the RW and trend baselines anchor forecasts to recent data but only the trend baseline incorporates the local trend of the time series. The model is fit to the log-transformed data (logit for ILI) but down-weights older observations exponentially, so that the estimated trend reflects recent dynamics. The damping parameter forces the median forecast to eventually flatten, rather than extrapolating the current trend indefinitely. All three baseline forecasts were constructed using versioned data that were available at the time of each actual forecast. While they were created retrospectively, they use the same data available for each forecast submitted in real time and could be produced as real-time forecasts. More details on each benchmark model are reported in the Methods.

Fig. 1A illustrates hospitalization forecasts for each benchmark model and the FluSight ensemble for five forecast dates in the 2023-2024 influenza season. We calculated the WIS for the unweighted ensembles from each challenge and each benchmark forecast on the logit (ILI) or log (others) scale [6], finding substantial variability across outcomes, seasons, and locations (Fig. 1B-F). Scores for the baseline forecasts varied by season, reflecting changes in observational noise, complexity of trajectories, and novelty, such as for COVID-19 cases or deaths in 2020 (no historical data), for daily COVID-19 hospitalizations in 2023 (the lowest recorded peak to date), or influenza hospitalizations in 2024-2025 (the highest recorded peak for the NHSN reporting system to date). Among the benchmark models, the RW and trend baseline models outperformed the naïve model in 92.9% and 95.9% of location-season-outcome combinations across 1-4 week horizons (748 and 772 of 805), respectively. Instances where the naïve baseline performed better were distributed across locations and outcomes, most notably for influenza hospitalizations in 2021-2022 and for COVID-19 deaths and hospitalizations across seasons.

**Figure 1:**
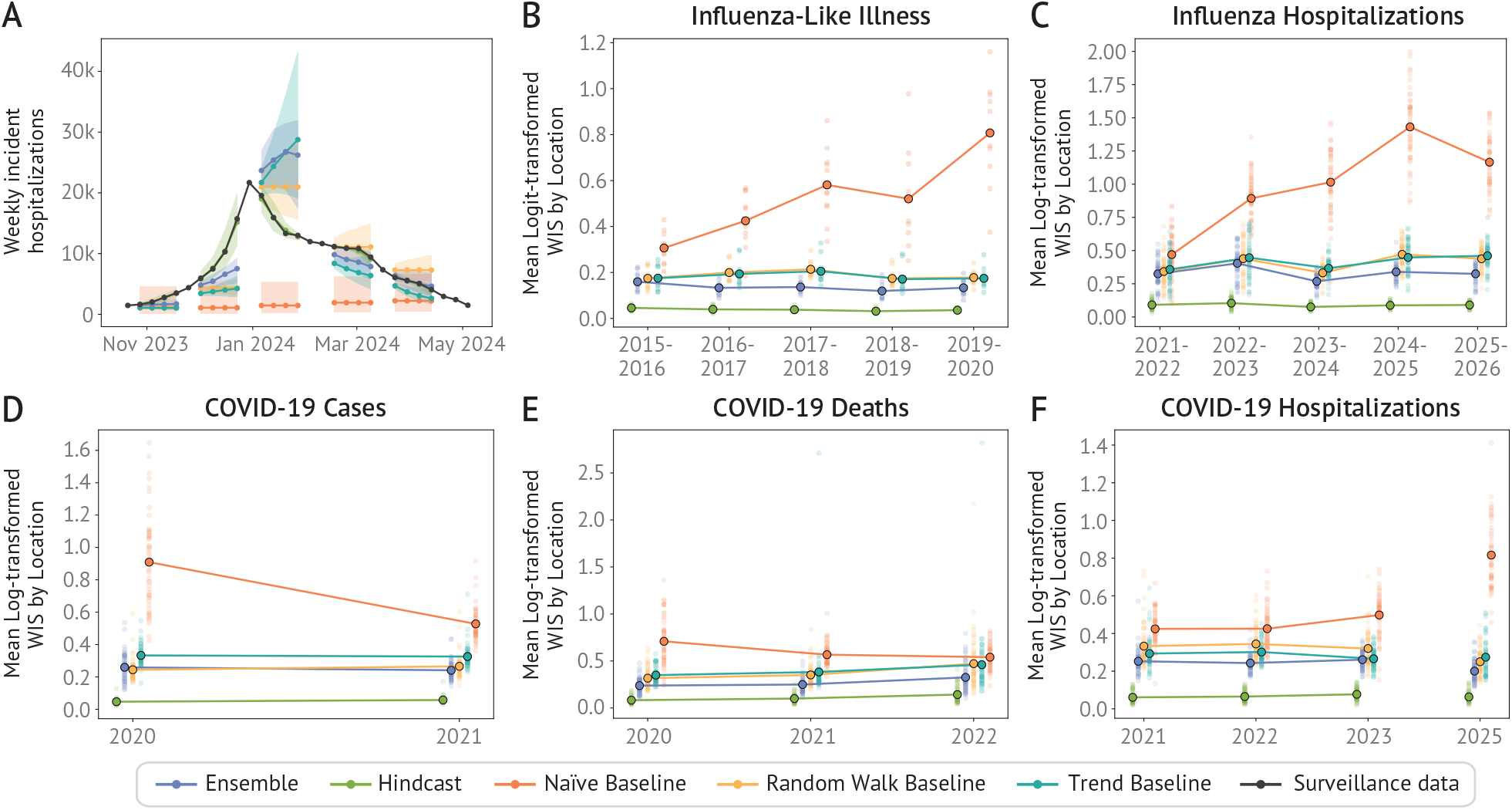
Example forecasts and proper scores for ensemble, optimal, and reference forecasts across influenza and COVID-19 forecasting targets and seasons. (A) Example of forecasts for the ensemble, optimal (the hindcast), and reference models (here, the three baseline models: naïve, random walk and trend) for the 2023-24 FluSight hospitalization forecast season at the U.S. national level. (B-F) Mean log/logit-transformed WIS for each season and target disease outcome. Light circles represent mean WIS in each state, darker circle represents the mean WIS across all locations. Shown for each forecasting season for the ensemble, optimal (the hindcast), and reference models (naïve, random walk and trend baselines).

Comparison to the optimal hindcast provides a signal-to-noise ratio reflecting how much outcome variability there is beyond observation variability. A ratio of one relative to the naïve forecast suggests no information can be gained from forecasts, and a ratio of one relative to the trend or RW baseline indicates no benefit from more sophisticated models. No baseline forecast outperformed the hindcast in any location-season-outcome combination. However, two location-season-outcome combinations came close, with a naïve-to-hindcast WIS ratio less than 2: COVID-19 deaths in Florida in 2022 (when reporting switched to bi-weekly) and COVID-19 daily hospitalizations in the U.S. Virgin Islands in 2022 (309 sporadic hospitalizations).

Both the RW and trend models outperformed the original baseline models for all challenges including the ILI baseline model, which was based on historical seasonality rather than the most recent data (Fig. S5). Comparing to a variety of other proposed baseline models [53], we also found similar or improved performance by the RW and trend baseline models (Fig. S7).

The three baseline models differ in their use of recent data and therefore their skill across horizons. The RW and trend models initially outperform the naïve forecast, but their performance degrades rapidly as the horizon increases (Fig. 2). In contrast, the naïve model is largely insensitive to horizon since it draws on all historical data, with no weighting to recent observations. In most seasons, the RW and trend baseline WIS crossed the naïve WIS at a horizon of 4-9 weeks, indicating that recent data were most informative for forecasts within this window. However, in seasons with anomalous dynamics that were not captured well by the naïve forecast, this crossover did not occur.

**Figure 2:**
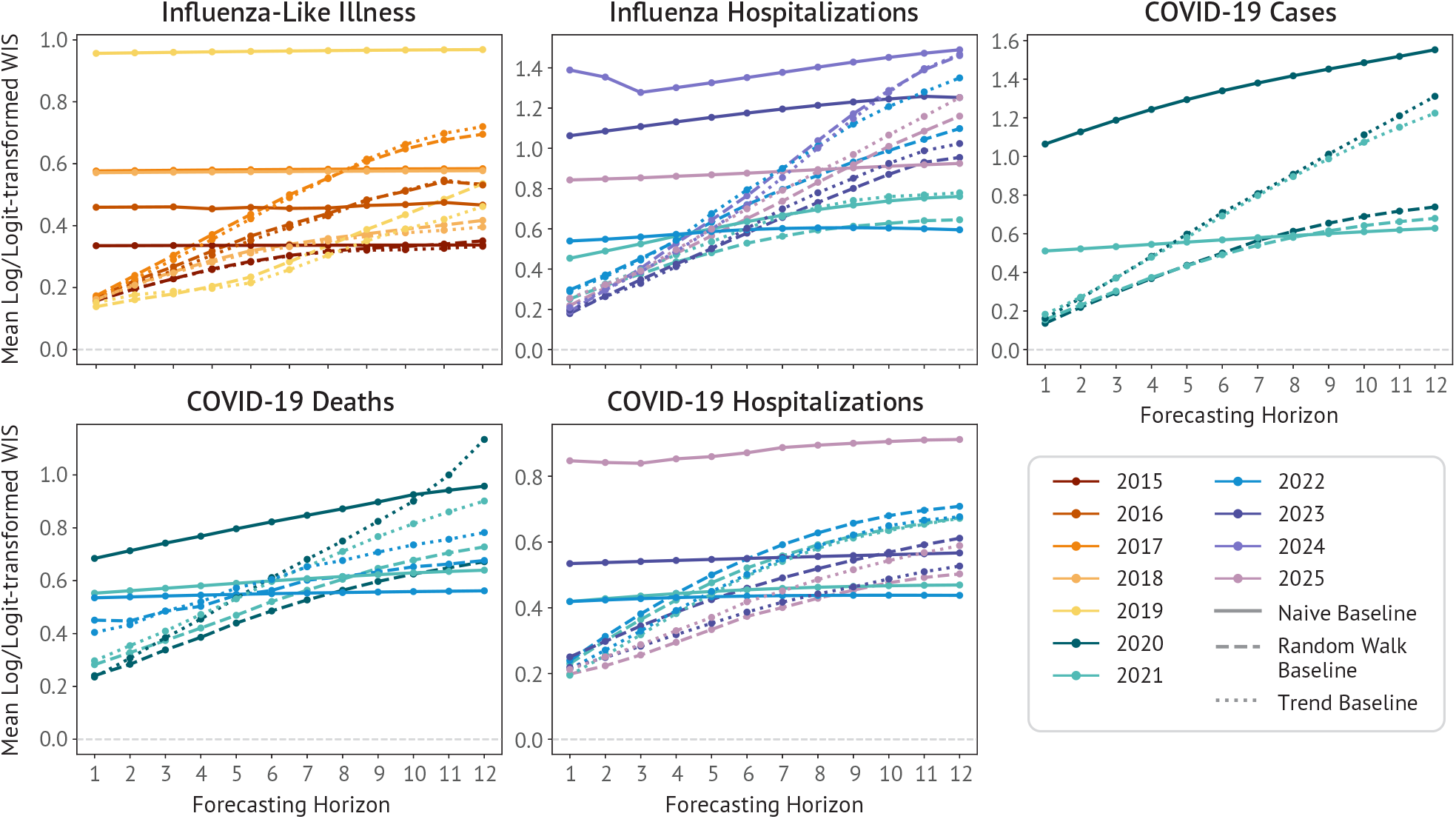
Baseline model performance for extended forecasting horizons by season and target. Mean log-transformed WIS for the 3 reference models at each horizon (up to 12 weeks ahead) and season for each target disease outcome. Solid lines show the score for the naïve model, dashed line the random walk baseline, and dotted line the trend baseline. Colors represent different seasons.

### Ensemble forecast skill

Across these seasons and outcomes, unweighted ensembles of all submitted forecasts represent the collective forecast effort of each challenge and were among the top forecasts in retrospective analyses [16, 33, 38, 46], so we first focused on assessing the performance of the mean or median ensemble forecasts for each challenge (Table S1). Since each baseline model captures a different aspect of a simplistic forecast, providing different performance bounds, comparison against all three offers a more rigorous test than any single baseline. Comparing the ensembles to the three baseline forecasts across 805 locations, outcomes, and seasons, the ensemble forecasts outperformed the RW baseline forecast for 90.7% of location-season-outcome combinations (730), the trend baseline for 90.3% (727), and the naïve forecasts for more than 98.9% (all but nine: 2021-2023 COVID-19 hospitalizations in the U.S. Virgin Islands, 2022 COVID-19 deaths in Alaska, and 2021-2022 influenza hospitalizations in Georgia, Mississippi, Tennessee, Virginia, and Wyoming, Fig. 1B–F). While the influenza hospitalization forecasts in 2021-2022 may represent chance outcomes in a new surveillance system during a season with atypical timing and low severity [14], the COVID-19 outcomes in the U.S. Virgin Islands and Alaska represent the challenge of forecasting when counts are low and the observation noise is similar to the signal in the data.

Across locations, the ensemble performed better than each baseline for all targets, seasons, and horizons except for three, the RW baseline for ILI at Horizon 1 in 2015-2016 and COVID-19 cases at Horizons 3 and 4 in 2020 (Fig. 3B). For all targets and horizons, skill scores were lower with the RW and trend models compared to the naïve, indicating that outperforming these baselines is generally a stricter test of model skill. However, the horizon specific effects of the RW and trend baseline skill scores show a counterintuitive effect. Visual inspection and proper scores show that forecast performance decreases at longer horizons (Fig. S2), yet for all ILI forecasts and some other outcomes and seasons, skill scores relative to the RW and trend baselines improved at longer forecast horizons (Fig. 3B & C). This results from changing horizon-specific performance in those baseline models (Fig. 2). At longer horizons, performance degrades faster for the RW and trend baselines than for the ensemble, so the skill score of the ensemble, a relative measure, increases even though actual forecast performance decreases. Therefore skill scores using RW and trend baselines, while valuable comparators for these baselines, do not provide reliable indicators of absolute performance across horizons. To analyze skill across horizons, a baseline with consistent performance across horizons, like the naïve baseline model is necessary. Skill scores using the naïve baseline model showed decreased accuracy at longer horizons, as expected, across all outcomes and seasons (Fig. 3A), contrasting the behavior in the RW and trend skill scores (Fig. S3).

**Figure 3:**
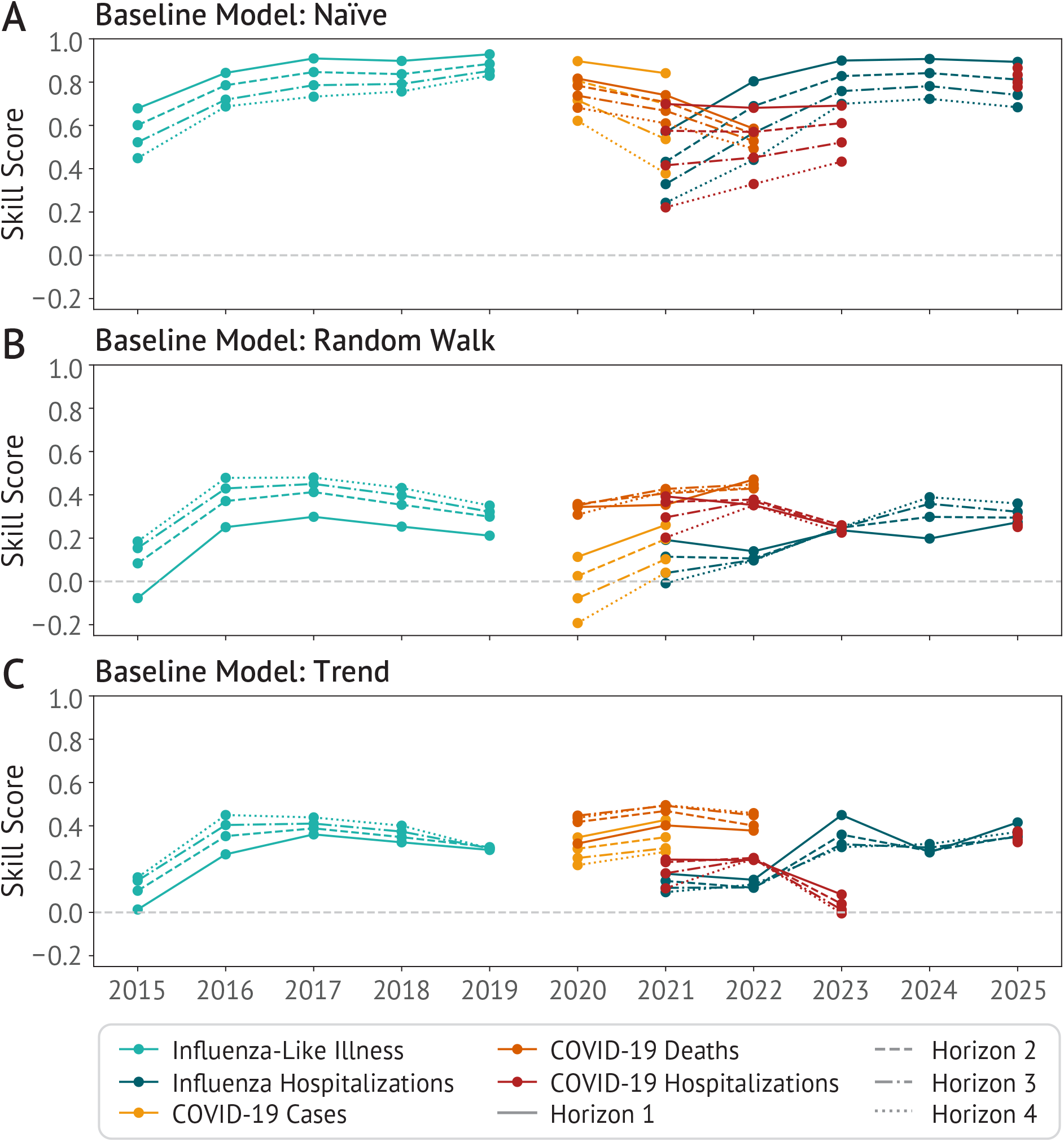
Ensemble skill score by season, target, and forecasting horizon relative to three baseline models. Skill score across season for each disease outcome for the ensemble reported to each forecast hub, separated by horizon, the number of weeks ahead predicted. The horizontal gray line represents a skill of zero, which is equivalent to the skill of the reference model. The reference model is (A) the naïve baseline, (B) the random walk baseline, and (C) the trend baseline.

The naïve baseline is critical to assess performance across horizons, but it is not adequate to assess skill at short-term horizons where it performs poorly. The RW and trend baselines provide more rigorous tests. For example, some of the ensemble forecasts (e.g., influenza hospitalizations at Horizon 1) appeared to approach a skill score of 1 relative to the naïve model, but most only showed marginal improvement over the RW and trend baselines indicating that the ensembles are still not approaching the skill of the hindcast. Each baseline model captures a different aspect of a simplistic forecast, providing different performance bounds, comparison against all three offers a more rigorous test than any single baseline. A forecast that outperforms all three provides stronger evidence of forecast skill beyond extrapolation of recent or historical patterns. The findings are consistent with relative WIS comparisons but provide a more reasonable upper bound of forecast performance which reflects outcome-specific variability (Fig. S4).

### Forecast improvement

For ILI, influenza hospitalizations, and COVID-19 daily hospitalizations ensembles, we found improved skill over time compared to the naïve baseline (Fig. 3A), with a plateau for influenza hospitalization forecast skill in 2024-2025 and 2025-2026. In contrast, the skill of ensemble forecasts for COVID-19 cases and deaths declined relative to the naïve model. The weekly COVID-19 hospitalization ensemble forecasts for 2025 showed high skill, but comparison across years was not possible due to that being the first year of weekly forecasts. The skill scores with the naïve baseline also enable horizon-specific analyses and, while skill uniformly decreased at longer horizons, it also changed over time. Comparing horizon-specific skill between the first and last season forecasted for each target, we found that skill improved for both the 1- and 4-week-ahead horizon across all locations for ILI and influenza hospitalizations (Fig. 4). Additionally, the 4-week-ahead forecasts for the most recent year had higher skill than the 1-week-ahead forecasts for the first year of the corresponding forecast challenge. For COVID-19, the results were mixed, as case and death forecast skill decreased and daily hospitalization forecast skill was relatively constant at the 1-week horizon while improving at the 4-week horizon, averaged across all locations (Fig. 4). The COVID-19 results also reflect the particular challenge of developing forecasts in the absence of historical data. The naïve baseline performs worst with novel observations, such as during emergence or surveillance changes. High skill relative to the naïve baseline in seasons with limited historical data can therefore reflect skill under those circumstances, but similar skill may be difficult to attain for a similar season once more data have accumulated.

**Figure 4:**
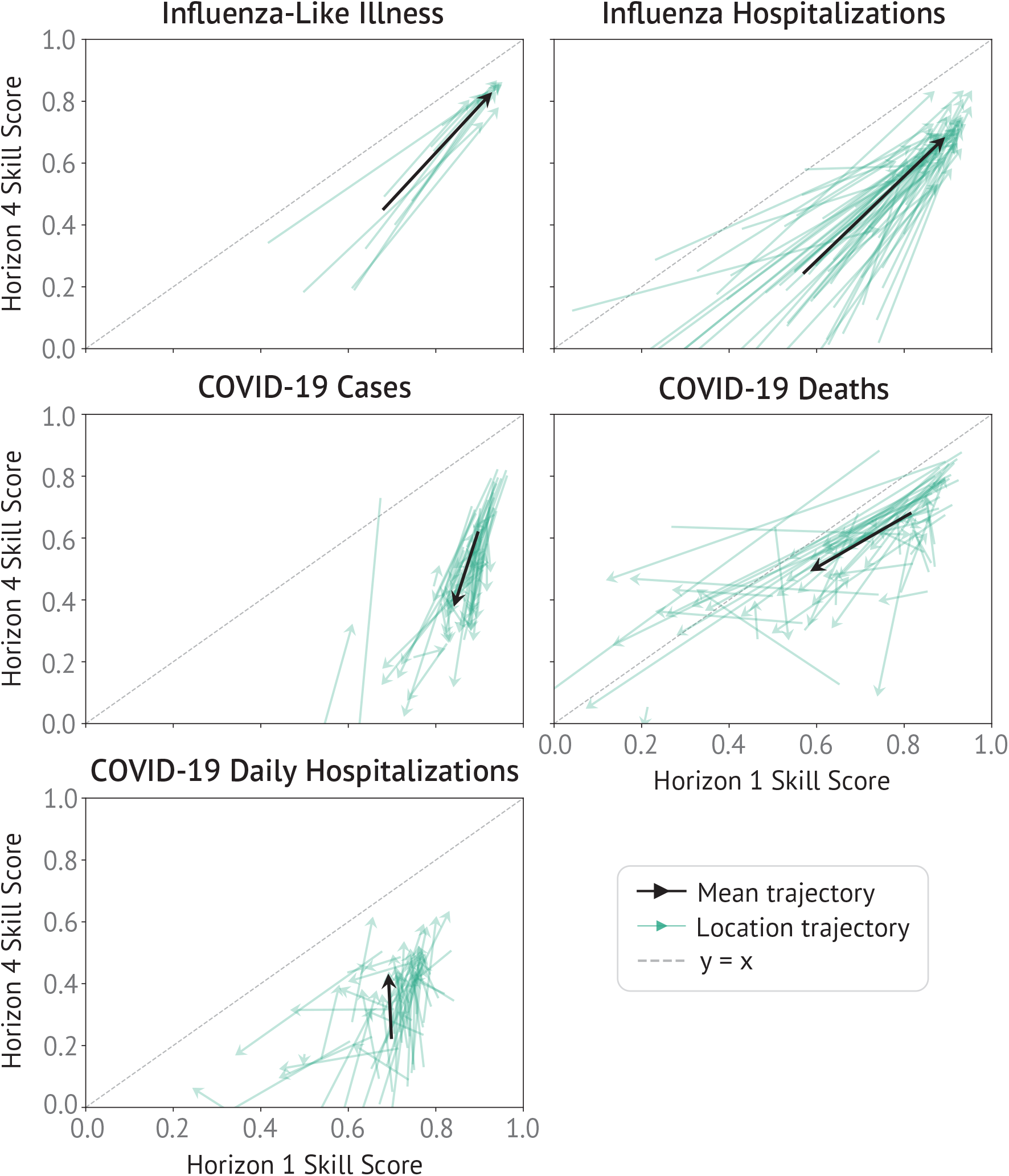
Horizon-specific changes in ensemble skill scores across seasons. Change in horizon 1 vs horizon 4 skill over time for each disease outcome and location. Green arrows show the change in horizon 1 vs. 4 skill for each location from the first (arrow start) to the last (or most recent) season (arrow end) forecasted. Arrow length represents the magnitude of the change in skill. For COVID-19 hospitalizations, we show daily forecast skill change for 2021-2023. The black arrows show the change in the overall skill across all locations. Skill score is computed relative to the naïve baseline.

Comparison to the RW and trend baselines provides complementary insights. In general, skill scores were substantially lower, reflecting that these baseline forecasts also perform much better than the naïve model (Fig. 3B & C). Skill for the ILI ensemble increased initially, plateaued, and then decreased slightly in the last two seasons, corresponding to seasons when the RW and trend baselines performed relatively well (Fig. 1B). COVID-19 case and death ensemble forecast performance improved relative to the RW baseline model and were stable relative to the trend baseline. Hospitalization ensemble forecasts for both influenza and COVID-19 (daily) improved relative to the RW and trend baseline forecasts, with the exception of COVID-19 in 2023. Both the RW and trend baselines outperformed ensembles for specific target-season-horizon combinations. Ensemble skill scores were negative compared to the RW baseline for ILI in 2015-2016 at horizon 1, COVID-19 cases in 2020 at horizons 3 and 4, and influenza hospitalization in 2021-2022 at horizon 4, all initial years of their respective challenges. Ensemble skill was only negative relative to the trend baseline for COVID-19 hospitalizations in 2023 at horizon 4.

Analyzing team-specific forecast skill over time showed a growing proportion of individual models outperforming each reference model for both influenza targets, alongside increasing numbers of participating teams, likely contributing to improved ensemble skill (Fig. S8). COVID-19 results were mixed, with dynamic participation and inconsistent team performance relative to the reference models. The majority of team forecasts outperformed the naïve baseline model across all seasons and disease outcomes. Fewer individual teams outperformed the RW and trend baselines, although the proportion of teams performing better than these baselines improved over time.

## Discussion

The field of infectious disease forecasting has grown significantly over the last decade, spurred in part by standardized real-time forecasting frameworks and evaluation. However, many important questions remain including: How good are the forecasts? And, have forecasts improved? Here, we addressed these questions using an expanded skill score framework with four new benchmark models and an analysis of over 2.5 million influenza and COVID-19 forecasts.

### Forecast benchmarks

We developed four specific benchmark forecasting models applied to COVID-19 and influenza. All are straightforward to implement using routine epidemiological data and require limited assumptions. More important than the specific model formulations are their distinctions in performance and function. The optimal hindcast model provides an improved target for forecasting skill with noisy, imperfectly observed outcomes. Baseline models using limited data to make reasonable short-term predictions have been used as benchmarks for infectious disease forecasting since 2015 [4, 30], and their design influences the interpretation of forecast performance [54]. Here, we implemented three baseline forecasts: a random walk forecast, a trend forecast, and a naïve forecast, each using data available to real-time forecasts. The RW baseline outperformed the original baseline models for each of the challenges analyzed here and alternative approaches proposed by others [53]. The trend baseline, despite incorporating recent trends, performed slightly worse. The naïve baseline is an even simpler model, similar to the baseline model used for the West Nile Virus forecasting challenge [21], a horizon-independent indicator of how unique observed outcomes are compared to past outcomes. It therefore allows assessment of relative forecast performance for common and uncommon observations, the latter being one of the key challenges for infectious disease forecasting, and can give valuable insight on skill across horizons. Together, these benchmark models provide a comprehensive framework for evaluating and interpreting forecast performance and can be easily adapted to future forecasting challenges.

### Are forecasts “good”?

For infectious disease forecasts to be useful, they should provide improved information on current levels of transmission (which our naïve baseline forecasts cannot), improved trend information beyond a simple baseline forecast, and, ideally, approach precision similar to the intrinsic noise in the epidemiological data. Comparing the ensemble forecasts for influenza and COVID-19 to three baseline models and an optimal hindcast, we found that these forecasts have provided valuable information on the pathogens and outcomes assessed here, yet still have ample room for improvement. Most team forecasts also outperformed the naïve forecasts but many performed worse than the random walk (RW) and trend baseline forecasts implemented here (Fig. S8). These findings are in line with the pairwise relative WIS that has been reported using a lower skill baseline forecast in COVID-19 and influenza forecasting analyses [16, 37], a relative metric that also gives counterintuitive horizon-specific behavior (Fig. S6).

While the skill scores show value in the forecasts, they also show that skill is still limited in comparison to the optimal hindcast. The RW and trend baselines represent stricter performance bounds than the naïve model and the ensemble forecasts rarely reached a skill score of 0.5 against them, with the remaining gap toward 1 reflecting signal the ensembles are not capturing. This is consistent with visual inspection of the forecasts (e.g., Fig. 1A); many ensemble forecasts track incidence well yet the ensemble and hindcast often diverge in times of rapid change. While the hindcast represents a more reasonable bound for forecast skill than perfect forecasts, there is no guarantee that it is achievable. The hindcast itself is an estimate of what may have occurred, subject to its own data and structural limitations. Moreover, predictability does not only depend on uncertainty in the outcome data; the perfect model for transmission is unknown and even if it were, uncertainty in input data also generates forecast error particularly in non-linear systems with interactions across scales [34]. However, the optimal hindcast is one limit and it can identify outcomes where forecasting may not be useful. The lowest baseline-to-hindcast ratio we found was for COVID-19 hospitalizations in the Virgin Islands, the only place where the ensemble performed worse than the naïve baseline for all seasons. As this ratio is dependent only on the benchmark forecasts, when historical data are available, it can be used *a priori* to assess the potential value of developing more complex forecasts.

### Have forecasts improved?

The skill score provides a framework that enables adjustment for forecast difficulty. Noisier week-to-week observations are more difficult to predict with precision, which is reflected in worse hindcast scores. Seasons or outcomes that are atypical or have higher variability are also more difficult to predict and are reflected in worse scores for the naïve forecasts. Relative to these two forecasts, we found consistent improvements in skill scores for the ILI and influenza hospitalization targets, including horizon specific improvements and increasing proportions of teams submitting models that outperformed the reference forecasts.

COVID-19 forecasts generally performed better than all three baseline models and improved relative to the RW and trend baseline but had mixed improvement relative to the naïve forecast. For case and death forecasts, ensemble skill peaked in 2020 when there was little historical information available for each forecast, but had lower skill in 2021 despite more teams contributing. In 2021, case reporting changed substantially, likely contributing to a decline in skill for both case forecasts and death forecasts, which often relied on case data [16]. COVID-19 case forecasts were discontinued in late 2021 due to poor performance, and COVID-19 death forecasts in 2022 after substantial reporting changes complicated forecasting and evaluation. COVID-19 daily hospitalization forecasts, on the other hand, showed improved performance at horizons of 3-4 weeks relative to the naïve forecasts, likely in part reflecting a shift in forecast effort to a sustained and standardized national reporting system. A further shift in these data occurred in 2024, when reporting changed from daily to weekly [22, 55], but it is too early to assess improvement for the new weekly forecasts.

Multiple factors likely have contributed to improved forecast performance for ILI, influenza hospitalization, and COVID-19 hospitalization targets. First, forecasts require data. In contrast to COVID-19 case and death forecasts, ILI and hospitalization forecasts leveraged national surveillance systems with standardized reporting. Additional historical data also likely contributes; ILI forecasts benefited from almost two decades of historical data and hospitalization forecasts improved over time as additional data accrued. The influenza targets were the two longest running challenges analyzed here, giving teams more training data and time to refine models. For COVID-19, surveillance began in 2020, concurrent with initial forecasting efforts and most COVID-19 death forecasts soon used case data as a leading indicator [16]. However, increased at-home testing in late 2021 destabilized case data [43], decoupling case, hospitalization, and death data. This coincided with the discontinuation of case forecasts and the declining skill of death forecasts. The reporting frequency for deaths also changed substantially in 2022, with some states transitioning to intermittent rather than weekly reporting.

Other data sources may also be critical as forecasts are increasingly using ancillary data on related outcomes (e.g., emergency department visits [8]), behavior (e.g., mobility [50] or interaction patterns [32]), and online activity (e.g., internet searches [52]). Approaches to use these data vary, from augmenting sparse surveillance data [2, 39] to integrating multiple data streams [45]. Machine learning methods, high-performance computing, and other technologies have also advanced over this time period, yet the contribution of new and improved data and models is not directly measurable due to the evolving and diverse models used to generate forecasts. Sustained engagement facilitated by forecasting challenges, interest, and funding (e.g., Insight Net) likely has helped drive improvement by enabling teams to focus on specific forecasting problems with standardized inputs, outputs, and evaluation metrics which have facilitated participation via improved forecast frameworks and enabled the identification of areas in which models succeed or fail [23, 31, 48]. Previous studies have identified that teams that repeatedly participated in a forecasting challenge tend to perform better [23, 38]. We found that ensemble improvement was generally accompanied by improvement in contributing team forecasts as well.

### Conclusion

Proper scores have enabled robust comparison of infectious disease forecasts, which is essential to improving forecasts, yet beyond a perfect proper score they are not meaningful when comparing across different forecast targets. Comparing proper scores to those of standardized benchmarks using the skill score gives them meaning. Equivalence to the hindcast indicates that a forecast is as good as can be expected. Better performance than an informative baseline indicates that the forecast outperforms a simple forecast. And better or improved performance relative to the naïve forecast indicates robustness and improvement across horizons and targets of varying difficulty. Skill scores can be used with any proper score. They are not strictly proper on their own, yet with many observations they become approximately proper [40, 59]. Nor are skill scores a magic bullet to solve all evaluation questions, but they add important context while maintaining the advantages of proper scores. Here, we used this framework to demonstrate ensemble and team forecast improvement over a decade of infectious disease forecasting, progress driven by standardized forecasting initiatives and advances in data and forecasting models.

## Methods

### Target Data

Surveillance data from multiple data sources were used as ground truth to evaluate forecasts across the six outcome targets. ILI data were reported through the ILINet System for U.S. HHS Regions and nationally [56]. Forecasts were evaluated against weighted ILI values, the proportion of outpatient visits due to influenza-like illness weighted for population size [38]. Influenza and weekly COVID-19 hospital admissions data were reported through the National Healthcare Safety Network (NHSN), which reports information on hospitalization capacity at the U.S. state and national level [55]. COVID-19 case and death data were reported by the Johns Hopkins CSSE group and consisted of city, state, and national level reports in the U.S. and globally [18, 19]. Here, we focused on the state and national counts in the United States. NHSN data were used as the ground truth for daily COVID-19 hospitalizations, with daily updates on confirmed hospital admissions at the state level [22].

### Weighted Interval Score

The weighted interval score (WIS) has become widely accepted for assessing forecasting predictions given in a quantile format, where smaller scores indicate improved performance [25, 7]. The WIS assesses quantile forecasts by analyzing the sharpness, or uncertainty in the predictions, as well as the calibration (agreement) of the predictions with respect to the observed data [25]. The WIS is computed at each time point with prediction *P* and observed value *y* as a weighted sum of the interval score for each prediction interval *α* using the expression

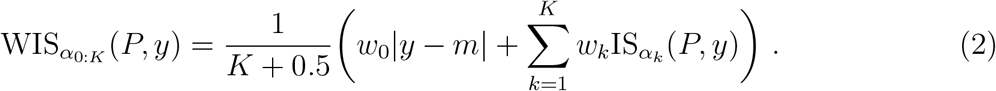

IS_*α*_ corresponds to the interval score of the prediction interval *α*,

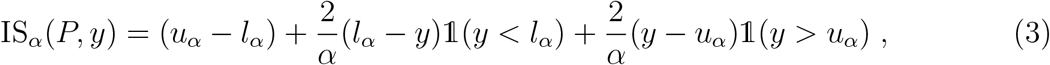

where *K* is the number of prediction intervals used, *m* is the median of *P*, and *l*_*α*_ (*u*_*α*_) is the lower bound (upper bound) of the prediction interval. The standard weights are chosen such that 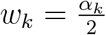 and 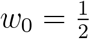 [7].

The WIS is dependent on the magnitude of the projection target across time and location. This constrains model comparison with the WIS for surveillance data with different orders of magnitude. To reduce this impact, we transformed the forecast and surveillance data to the logarithmic scale and computed the WIS on the log-transformed values [6], adding 1,log(*x*+1), to avoid log(0) issues. For ILI forecasts, which are reported on the percent scale, we computed the WIS on logit-transformed scales after converting the binned probability distributions to quantiles via interpolation (see Supplementary Material).

### Optimal Hindcast Model

For the optimal model, we generated a hindcast, which differs from a traditional forecast by providing a probabilistic interpretation of the surveillance data, adjusting for noise in the observations rather than predicting future trends. This treats the observed outcome as a noisy observation from a time series rather than as ‘truth.’ This is a retrospective model that mimics the performance of what we might consider an optimal, or perfect, forecast. Although hindcasts likely overestimates attainable forecast skill since it incorporates data unavailable for prospective forecasts [51], hindcasts are widely used in climate modeling for evaluating relative performance against forecasting models and identifying model parameters [36]. In this work, the hindcast fits the mean and dispersion of a 3-week moving window (the current week and its immediate neighbors) of the epidemiological time series to a negative binomial distribution (logit-normal for ILI percentages). Extra weight (double) is given to the center (current) week. The fitted parameters capture both the local level and variability of the observed signal at each location and time point. The hindcast is static for a given date (regardless of forecast date).

### Reference Model

For the reference model, we consider three types of baseline forecasts that should generally be uninformative and consistently producible [24]. These serve as a useful benchmark for comparison, as it allows us to assess the added value and accuracy of models using more sophisticated, data- or mechanism driven methods [44]. The baseline models adjust for difficulty in forecasting, e.g., compared to past observations. To ensure fair comparison with forecasts generated in real time, we generated the baseline forecasts using versioned surveillance data compiled by the Delphi Epidata API, which tracks historical data revisions for a variety of epidemic datasets [50].

We generated a naïve, random walk, and trend forecast as the three baseline models. The naïve forecast is based only on historical observations, without conditioning on recent data. This framework is a common choice in weather and climate forecasting [60]. The model is produced by fitting a negative binomial distribution to all available historical observations at a given location prior to the forecast date, capturing the overall level and variability of historically observed data. This distribution is applied identically across forecast horizons, producing flat uncertainty for future horizons. This naïve model represents long-term behavior, and does not capture seasonal trends, or periods of increased activity.

For the random walk (RW) baseline, we used a log-transformed version of the baseline method used by the COVID-19 Forecast Hub, and FluSight Forecast Hub since 2021 [44]. This method generates flat forecasts, where the median for each week ahead is equivalent to the most recently observed data point with expanding uncertainty for longer horizons. To do this, weekly differences between consecutive weeks of surveillance data are sampled and added to the previous weeks’ samples for longer horizons [44]. The weekly differences are symmetrized to give an equal probability of observing an increase or decrease at future horizons. These differences are then added to the most recent surveillance data. Here, we log-transformed the surveillance data (log(*x* + 1)) before generating the RW baseline, and then re-transformed the resulting forecast. For the FluSight ILI forecasts, we performed the same method using the logit transform. While this model projects a flat median, the forecast is entirely uncertain about whether future observations with increase or decrease; it is essentially a random walk on historically observed week-to-week differences.

The trend baseline is an exponential smoothing model with additive errors, a damped additive trend, and no seasonality, ETS(*A, A*_*d*_, *N*), according to the notation of [26]. In the innovations state space form, the model is written as

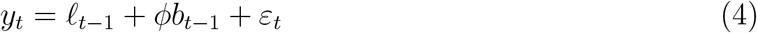

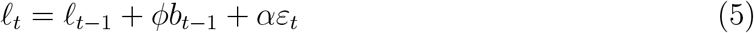

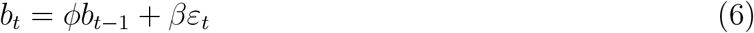

where *l*_*t*_ is the level, *b*_*t*_ is the trend, and *ε*_*t*_ is an independent normally distributed error with mean zero and variance *σ*^2^. The smoothing parameters *α* and *β* control the rate at which the level and trend adapt to new observations, respectively. The damping parameter *ϕ* ∈ (0, 1) causes the trend to decay geometrically across forecast horizons, so that the *h*-step ahead point forecast is

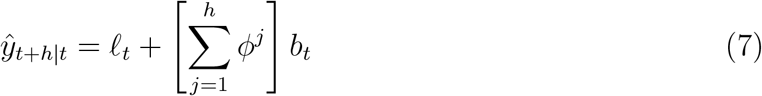

As *h → ∞*, point forecasts converge to the finite limit 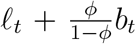. The smoothing parameters *α, β, ϕ* and initial states *l*_0_ and *b*_0_ are estimated by maximum likelihood, and prediction intervals are derived from the *h*-step ahead forecast error variance, assuming normally distributed errors. This model was fitted to the log transformed surveillance data (or logit transformed in the case of ILI), using the forecast package in R [27, 29], with training data restricted to observations available at each forecast date. For more details, see [28] and Chapter 7 of [26].

## Data Availability

All data produced are available online at

https://doi.org/10.5281/zenodo.23047785

## Data and Code Availability

Our code for this project is publicly available on Zenodo [1].

## Acknowledgments

We acknowledge support from the cooperative agreement CDC-RFA-FT-23-0069 from the CDC’s Center for Forecasting and Outbreak Analytics.

## Disclaimer

The findings and conclusions in this study are those of the authors and do not necessarily represent the official position of the funding agencies, the CDC, or the U.S. Department of Health and Human Services of the United States.

## References

[1] Bay, C., 2026. clarabay/skill-score: Release v1.0. URL: https://doi.org/10.5281/zenodo.23047786, doi:10.5281/zenodo.23047786.

[2] Benefield, A.E., Williams, D., Nagraj, V., 2025. An imputation-based approach for augmenting sparse epidemiological signals. medRxiv URL: https://www.medrxiv.org/content/early/2025/07/29/2024.07.31.24311314, doi:10.1101/2024.07.31.24311314.

[3] Biggerstaff, M., Alper, D., Dredze, M., Fox, S., Fung, I.C.H., Hickmann, K.S., Lewis, B., Rosenfeld, R., Shaman, J., Tsou, M.H., Velardi, P., Vespignani, A., Finelli, L., for the Influenza Forecasting Contest Working Group, 2016. Results from the centers for disease control and prevention’s predict the 2013–2014 Influenza Season Challenge. BMC Infectious Diseases 16, 357. URL: https://doi.org/10.1186/s12879-016-1669-x, doi:10.1186/s12879-016-1669-x.

[4] Biggerstaff, M., Johansson, M., Alper, D., Brooks, L.C., Chakraborty, P., Farrow, D.C., Hyun, S., Kandula, S., McGowan, C., Ramakrishnan, N., Rosenfeld, R., Shaman, J., Tibshirani, R., Tibshirani, R.J., Vespignani, A., Yang, W., Zhang, Q., Reed, C., 2018. Results from the second year of a collaborative effort to forecast influenza seasons in the United States. Epidemics 24, 26–33. URL: https://www.sciencedirect.com/science/article/pii/S1755436517300889, doi:10.1016/j.epidem.2018.02.003.

[5] Biggerstaff, M., Slayton, R.B., Johansson, M.A., Butler, J.C., 2022. Improving Pandemic Response: Employing Mathematical Modeling to Confront Coronavirus Disease 2019. Clin. Infect. Dis. 74, 913–917. URL: https://doi.org/10.1093/cid/ciab673, doi:10.1093/cid/ciab673.

[6] Bosse, N.I., Abbott, S., Cori, A., van Leeuwen, E., Bracher, J., Funk, S., 2023. Scoring epidemiological forecasts on transformed scales. PLOS Computational Biology 19, 1– 23. URL: https://doi.org/10.1371/journal.pcbi.1011393, doi:10.1371/journal.pcbi.1011393.

[7] Bracher, J., Ray, E.L., Gneiting, T., Reich, N.G., 2021. Evaluating epidemic forecasts in an interval format. PLOS Computational Biology 17, 1–15. URL: https://doi.org/10.1371/journal.pcbi.1008618, doi:10.1371/journal.pcbi.1008618.

[8] CDC, U., 2026. National Syndromic Surveillance Program - About NSSP. https://www.cdc.gov/nssp/php/about/index.html. Accessed: 2026-05-01.

[9] Centers for Disease Control and Prevention, 2023. FluSight Forecast Data. https://github.com/cdcepi/Flusight-forecast-data. Accessed: 2026-05-22.

[10] Centers for Disease Control and Prevention, 2024. Epidemic Prediction Initiative. https://archive.cdc.gov/www_cdc_gov/epidemic-prediction-initiative/index_1723639804.html. Accessed: 2025-02-13.

[11] Centers for Disease Control and Prevention, 2025. COVID-19 Forecast Hub. https://github.com/CDCgov/covid19-forecast-hub/. Accessed: 2025-10-17.

[12] Centers for Disease Control and Prevention, 2026a. FluSight Forecast Hub. https://github.com/cdcepi/FluSight-forecast-hub/tree/main. Accessed: 2026-05-22.

[13] Centers for Disease Control and Prevention, 2026b. FluSight Forecasts. https://github.com/cdcepi/FluSight-forecasts. Accessed: 2026-05-22.

[14] Centers for Disease Control and Prevention, 2026c. Past Flu Season Severity Assessments. https://www.cdc.gov/flu/php/surveillance/past-seasons.html. Accessed: 2026-08-18.

[15] Christen, P., Ombajo, L.A., Cori, A., Dawa, J., Djaafara, B.A., Kimani, T.N., Schneider, C.M.J., van Elsland, S.L., Thumbi, M., Veras, M., Whittaker, C., Whittles, L.K., Watson, O.J., 2026. Enhancing epidemic forecast usability for policymakers: A global mixed-methods study. PLOS Global Public Health 6, 1–17. URL: https://doi.org/10.1371/journal.pgph.0006519, doi:10.1371/journal.pgph.0006519.

[16] Cramer, E.Y., et al., 2022a. Evaluation of individual and ensemble probabilistic forecasts of covid-19 mortality in the united states. Proc. Natl. Acad. Sci. U.S.A. 119, e2113561119. doi:10.1073/pnas.2113561119.

[17] Cramer, E.Y., et al., 2022b. The united states COVID-19 forecast hub dataset. Scientific Data 9, 462. doi:10.1038/s41597-022-01517-w.

[18] Dong, E., Du, H., Gardner, L., 2020. An interactive web-based dashboard to track COVID-19 in real time. Lancet Infect. Dis. 20, 533–534. doi:10.1016/S1473-3099(20)30120-1.

[19] Dong, E., Ratcliff, J., Goyea, T.D., Katz, A., Lau, R., Ng, T.K., Garcia, B., Bolt, E., Prata, S., Zhang, D., Murray, R.C., Blake, M.R., Du, H., Ganjkhanloo, F., Ahmadi, F., Williams, J., Choudhury, S., Gardner, L.M., 2022. The Johns Hopkins University Center for Systems Science and Engineering COVID-19 Dashboard: data collection process, challenges faced, and lessons learned. Lancet Inf. Dis. 22, e370–e376. doi:10.1016/S1473-3099(22)00434-0.

[20] Gneiting, T., Raftery, A.E., 2007. Strictly proper scoring rules, prediction, and estimation. Journal of the American Statistical Association 102, 359–378. URL: https://doi.org/10.1198/016214506000001437, doi:10.1198/016214506000001437.

[21] Harp, R.D., Holcomb, K.M., Benjamin, S.G., Green, B.W., Jones, H., Johansson, M.A., 2026. A regionally determined climate-informed west nile virus forecast technique. GeoHealth 10, e2025GH001657. URL: https://agupubs.onlinelibrary.wiley.com/doi/abs/10.1029/2025GH001657, doi:10.1029/2025GH001657.

[22] HealthData.gov, 2025. COVID-19 Reported Patient Impact and Hospital Capacity by State Timeseries. https://healthdata.gov/Hospital/COVID-19-Reported-Patient-Impact-and-Hospital-Capa/g62h-syeh. Accessed: 2026-05-22.

[23] Hines, A.G., Mathis, S.M., Johansson, M.A., Biggerstaff, M., Reed, C., Borchering, R., 2026. A decade of the center for disease control and prevention’s flusight influenza forecasting. medRxiv URL: https://www.medrxiv.org/content/early/2026/06/08/2026.06.05.26354941, doi:10.64898/2026.06.05.26354941.

[24] Holcomb, K.M., Mathis, S., et al., 2023. Evaluation of an open forecasting challenge to assess skill of West Nile virus neuroinvasive disease prediction. Parasites & Vectors 16, 11. URL: https://doi.org/10.1186/s13071-022-05630-y, doi:10.1186/s13071-022-05630-y.

[25] Howerton, E., Contamin, L., et al., 2023. Evaluation of the US COVID-19 scenario modeling hub for informing pandemic response under uncertainty. Nature Communications 14, 7260. URL: https://doi.org/10.1038/s41467-023-42680-x, doi:10.1038/s41467-023-42680-x.

[26] Hyndman, R., Athanasopoulos, G., 2018. Forecasting: Principles and Practice. Second ed., OTexts: Melbourne, Australia. https://otexts.com/fpp2/ (Accessed 2026-05-22).

[27] Hyndman, R., Athanasopoulos, G., Bergmeir, C., Caceres, G., Chhay, L., O’Hara-Wild, M., Petropoulos, F., Razbash, S., Wang, E., Yasmeen, F., 2026. forecast: Forecasting functions for time series and linear models. URL: https://pkg.robjhyndman.com/forecast/, doi:10.32614/CRAN.package.forecast. r package version 9.0.2.

[28] Hyndman, R.J., 2008. Forecasting with exponential smoothing: the state space approach. Springer series in statistics, Springer, Berlin.

[29] Hyndman, R.J., Khandakar, Y., 2008. Automatic time series forecasting: the forecast package for R. Journal of Statistical Software 27, 1–22. doi:10.18637/jss.v027.i03.

[30] Johansson, M.A., et al., 2019. An open challenge to advance probabilistic forecasting for dengue epidemics. Proc. Natl. Acad. Sci. U.S.A. 116, 24268–24274. URL: https://www.pnas.org/doi/abs/10.1073/pnas.1909865116, doi:10.1073/pnas.1909865116.

[31] Kerr, M., Borchering, R., Rivadeneira, A.C., Contamin, L., Funk, S., Hochheiser, H., Howerton, E., Krystalli, A., Shandross, L., Reich, N.G., 2026. A software platform for collaborative infectious disease modelling. Nature Health, 1–10 URL: https://www.nature.com/articles/s44360-026-00145-7, doi:10.1038/s44360-026-00145-7.

[32] Litvinova, M., Sinclair, S., Kummer, A.G., Ventura, P.C., Foster, T., Shioda, K., Halloran, M.E., Vespignani, A., Ajelli, M., 2025. Epistorm-mix: Mapping social contact patterns for respiratory pathogen spread in the post-pandemic united states. medRxiv URL: https://www.medrxiv.org/content/early/2025/11/21/2025.11.20.25340662, doi:10.1101/2025.11.20.25340662.

[33] Lopez, V.K., Cramer, E.Y., et al., 2024. Challenges of covid-19 case forecasting in the us, 2020–2021. PLOS Computational Biology 20, 1–25. URL: https://doi.org/10.1371/journal.pcbi.1011200, doi:10.1371/journal.pcbi.1011200.

[34] Lorenz, E., 1969. How much better can weather prediction become? Technology Review 71, 38–49.

[35] Lutz, C.S., Huynh, M.P., Schroeder, M., Anyatonwu, S., Dahlgren, F.S., Danyluk, G., Fernandez, D., Greene, S.K., Kipshidze, N., Liu, L., et al., 2019. Applying infectious disease forecasting to public health: a path forward using influenza forecasting examples. BMC Public Health 19, 1–12.

[36] Ma, H.Y., Chuang, C.C., Klein, S.A., Lo, M.H., Zhang, Y., Xie, S., Zheng, X., Ma, P.L., Zhang, Y., Phillips, T.J., 2015. An improved hindcast approach for evaluation and diagnosis of physical processes in global climate models. Journal of Advances in Modeling Earth Systems 7, 1810–1827. URL: https://agupubs.onlinelibrary.wiley.com/doi/abs/10.1002/2015MS000490, doi:10.1002/2015MS000490.

[37] Mathis, S.M., Webber, A.E., et al., 2024. Evaluation of FluSight influenza forecasting in the 2021–22 and 2022–23 seasons with a new target laboratory-confirmed influenza hospitalizations. Nature Communications 15, 6289. URL: https://www.nature.com/articles/s41467-024-50601-9, doi:10.1038/s41467-024-50601-9.

[38] McGowan, C.J., Biggerstaff, M., Johansson, M., Apfeldorf, K.M., Ben-Nun, M., Brooks, L., Convertino, M., Erraguntla, M., Farrow, D.C., Freeze, J., Ghosh, S., Hyun, S., Kandula, S., Lega, J., Liu, Y., Michaud, N., Morita, H., Niemi, J., Ramakrishnan, N., Ray, E.L., Reich, N.G., Riley, P., Shaman, J., Tibshirani, R., Vespignani, A., Zhang, Q., Reed, C., 2019. Collaborative efforts to forecast seasonal influenza in the United States, 2015–2016. Scientific Reports 9, 683. URL: https://www.nature.com/articles/s41598-018-36361-9, doi:10.1038/s41598-018-36361-9.

[39] Meyer, A.G., Lu, F., Clemente, L., Santillana, M., 2025. A prospective real-time transfer learning approach to estimate influenza hospitalizations with limited data. Epidemics 50, 100816. URL: https://www.sciencedirect.com/science/article/pii/S1755436525000040, doi:10.1016/j.epidem.2025.100816.

[40] Murphy, A.H., 1973. Hedging and Skill Scores for Probability Forecasts URL: https://journals.ametsoc.org/view/journals/apme/12/1/1520-0450_1973_012_0215_hassfp_2_0_co_2.xml.

[41] Murphy, A.H., 1988. Skill Scores Based on the Mean Square Error and Their Relationships to the Correlation Coefficient URL: https://journals.ametsoc.org/view/journals/mwre/116/12/1520-0493_1988_116_2417_ssbotm_2_0_co_2.xml, doi:10.1175/1520-0493(1988)116<2417:SSBOTM>2.0.CO;2.

[42] Murphy, A.H., 1992. Climatology, persistence, and their linear combination as standards of reference in skill scores. Weather and Forecasting 7, 692 – 698. URL: https://journals.ametsoc.org/view/journals/wefo/7/4/1520-0434_1992_007_0692_cpatlc_2_0_co_2.xml, doi:10.1175/1520-0434(1992)007<0692:CPATLC>2.0.CO;2.

[43] Rader, B., Gertz, A., Iuliano, D., Gilmer, M., Wronski, L., Astley, C., Sewalk, K., Varrelman, T.J., Cohen, J., Parikh, R., Reese, H., Reed, C., Brownstein, J., 2022. Use of At-Home COVID-19 Tests — United States, August 23, 2021–March 12, 2022. MMWR Morb Mortal Wkly Rep 2022 71, 489–494. URL: https://www.cdc.gov/mmwr/volumes/71/wr/mm7113e1.htm?s_cid=mm7113e1_w, doi:10.15585/mmwr.mm7113e1.

[44] Ray, E.L., Brooks, L.C., Bien, J., Biggerstaff, M., Bosse, N.I., Bracher, J., Cramer, E.Y., Funk, S., Gerding, A., Johansson, M.A., Rumack, A., Wang, Y., Zorn, M., Tibshirani, R.J., Reich, N.G., 2023. Comparing trained and untrained probabilistic ensemble forecasts of covid-19 cases and deaths in the united states. International Journal of Forecasting 39, 1366–1383. URL: https://www.sciencedirect.com/science/article/pii/S0169207022000966, doi:10.1016/j.ijforecast.2022.06.005.

[45] Ray, E.L., Wang, Y., Wolfinger, R.D., Reich, N.G., 2025. Flusion: Integrating multiple data sources for accurate influenza predictions. Epidemics 50, 100810. URL: https://www.sciencedirect.com/science/article/pii/S1755436524000719, doi:10.1016/j.epidem.2024.100810.

[46] Reich, N.G., et al., 2019a. Accuracy of real-time multi-model ensemble forecasts for seasonal influenza in the U.S. PLOS Comput. Biol. 15, 1–19. URL: https://doi.org/10.1371/journal.pcbi.1007486, doi:10.1371/journal.pcbi.1007486.

[47] Reich, N.G., et al., 2019b. A collaborative multiyear, multimodel assessment of seasonal influenza forecasting in the United States. Proc. Natl. Acad. Sci. U.S.A. 116, 3146–3154. URL: https://www.pnas.org/doi/abs/10.1073/pnas.1812594116, doi:10.1073/pnas.1812594116.

[48] Reich, N.G., et al., 2022. Collaborative hubs: Making the most of predictive epidemic modeling. Am. J. Public Health 112, 839–842. URL: https://doi.org/10.2105/AJPH.2022.306831, doi:10.2105/AJPH.2022.306831.

[49] Reich Lab, 2024. COVID-19 Forecast Hub. https://github.com/reichlab/covid19-forecast-hub/tree/master. Accessed: 2026-05-22.

[50] Reinhart, A., Brooks, L., et al., 2021. An open repository of real-time covid-19 indicators. Proceedings of the National Academy of Sciences 118, e2111452118. URL: https://www.pnas.org/doi/abs/10.1073/pnas.2111452118, doi:10.1073/pnas.2111452118.

[51] Risbey, J.S., Squire, D.T., Black, A.S., DelSole, T., Lepore, C., Matear, R.J., Monselesan, D.P., Moore, T.S., Richardson, D., Schepen, A., Tippett, M.K., Tozer, C.R., 2021. Standard assessments of climate forecast skill can be misleading. Nature Communications 12, 4346. URL: https://www.nature.com/articles/s41467-021-23771-z, doi:10.1038/s41467-021-23771-z.

[52] Santillana, M., Nguyen, A.T., Dredze, M., Paul, M.J., Nsoesie, E.O., Brownstein, J.S., 2015. Combining search, social media, and traditional data sources to improve influenza surveillance. PLOS Computational Biology 11, 1–15. URL: https://doi.org/10.1371/journal.pcbi.1004513, doi:10.1371/journal.pcbi.1004513.

[53] Stapper, M., Funk, S., 2025. Mind the baseline: The hidden impact of reference model selection on forecast assessment. medRxiv URL: https://www.medrxiv.org/content/early/2025/09/03/2025.08.01.25332807, doi:10.1101/2025.08.01.25332807, arXiv:https://www.medrxiv.org/content/early/2025/09/03/2025.08.01.25332807.full.pdf.

[54] Suez, E., Fox, S.J., 2026. Basic baseline model design choices can substantially influence performance in collaborative forecast hubs. medRxiv URL: https://www.medrxiv.org/content/early/2026/03/20/2026.03.18.26348748, doi:10.64898/2026.03.18.26348748.

[55] US Centers for Disease Control and Prevention, 2025. National Healthcare Safety Network (NHSN). https://www.cdc.gov/nhsn/psc/hospital-respiratory-reporting.html. Accessed: 2026-05-22.

[56] US Centers for Disease Control and Prevention, 2026. FluView Interactive: National, Regional, and State Level Outpatient Illness and Viral Surveillance. https://gis.cdc.gov/grasp/fluview/fluportaldashboard.html. Accessed: 2026-05-21.

[57] Viboud, C., Vespignani, A., 2019. The future of influenza forecasts. Proceedings of the National Academy of Sciences 116, 2802–2804. URL: https://www.pnas.org/doi/abs/10.1073/pnas.1822167116, doi:10.1073/pnas.1822167116.

[58] Viboud, C., et al., 2018. The RAPIDD ebola forecasting challenge: Synthesis and lessons learnt. Epidemics 22, 13–21. URL: https://www.sciencedirect.com/science/article/pii/S1755436517301275, doi:10.1016/j.epidem.2017.08.002. the RAPIDD Ebola Forecasting Challenge.

[59] Wheatcroft, E., 2019. Interpreting the skill score form of forecast performance metrics. International Journal of Forecasting 35, 573–579. URL: https://www.sciencedirect.com/science/article/pii/S0169207019300093, doi:10.1016/j.ijforecast.2018.11.010.

[60] Wilks, D.S., 2006. Statistical Methods in the Atmospheric Sciences. Second ed., Elsevier.

