## Supplementary Material for "Quantifying advances in infectious disease forecast skill across seasons and pathogens"

#### Contents

**1 Ensemble Models** **1**

**2 Logarithmic Score** **2**

**3 Forecast Format Sensitivity Analysis** **3**

**4 Additional Horizon-Specific Analyses** **5**

**5 Impact of the Hindcast on Forecast Skill** **5**

**6 Submitted Baseline Comparison** **6**

**7 Baseline Model Selection** **9**

**8 Team Forecasts** **10**

#### 1 Ensemble Models

In the analysis presented here, we focus on the performance of the ensemble model produced by each forecasting hub. After receiving submissions from individual modeling teams, the coordination team of each forecast hub generate a multi-model ensemble forecast by aggregating the results from

Table S1: **Description of the ensemble model for each forecasting challenge considered in our analysis.**

| **Disease Outcome** | **Ensemble Model** | **Generation Method** | **Median no. of weekly submitted models** |
|---|---|---|---|
| Influenza-Like Illness | UnwghtAvg | Unweighted mean of submitted models' binned probabilities | 2015-16: 13, 2016-17: 25.5, 2017-18: 22, 2018-19: 33, 2019-20: 30 |
| Influenza Weekly Hospitalizations | FluSight-ensemble | Unweighted median of submitted models' quantiles. | 2021-22: 23, 2022-23: 21, 2023-24: 31.5, 2024-25: 41, 2025-26: 48 |
| COVID-19 Cases | COVIDhub-4_week_ensemble | Unweighted mean (before July 28, 2020)/ median (after July 28, 2020) of submitted models' quantiles | 2020: 12, 2021: 17.5 |
| COVID-19 Deaths | COVIDhub-4_week_ensemble | Unweighted mean (before July 28, 2020) / median (after July 28, 2020) of submitted models' quantiles | 2020: 16, 2021: 20, 2022: 14 |
| COVID-19 Daily Hospitalizations | COVIDhub-4_week_ensemble | Unweighted median of submitted models' quantiles | 2021: 5, 2022: 11, 2023: 10 |
| COVID-19 Weekly Hospitalizations | CovidHub-ensemble | Unweighted median of submitted models' quantiles. | 2025: 13.5 |

individual modeling teams into a single prediction [1]. We describe the ensemble model for each forecasting challenge in Table S1. Individual team forecasts, which vary in forecasting methodology, weeks submitted, and locations/horizons predicted, are also evaluated for each target outcome. We analyze the forecast dates corresponding to when the ensemble model was reported for each target outcome, which typically occurred on a weekly basis during each forecasting season.

### 2 Logarithmic Score

For forecasts submitted as binned predictive distributions, the original evaluations used the logarithmic, or log, score. The log score (LogS) is a proper score computed for a given prediction $P$ with probability $p_y$ given to the observed value $y$, and is defined as:

$$\mathrm{LogS(P, y) = \log p_y}, \quad (1)$$

where log is the natural logarithm [2, 3]. For consistency and ease of understanding, we reported the negative log score (-LogS) such that smaller scores represent a higher probability placed on the true outcome.

### 3 Forecast Format Sensitivity Analysis

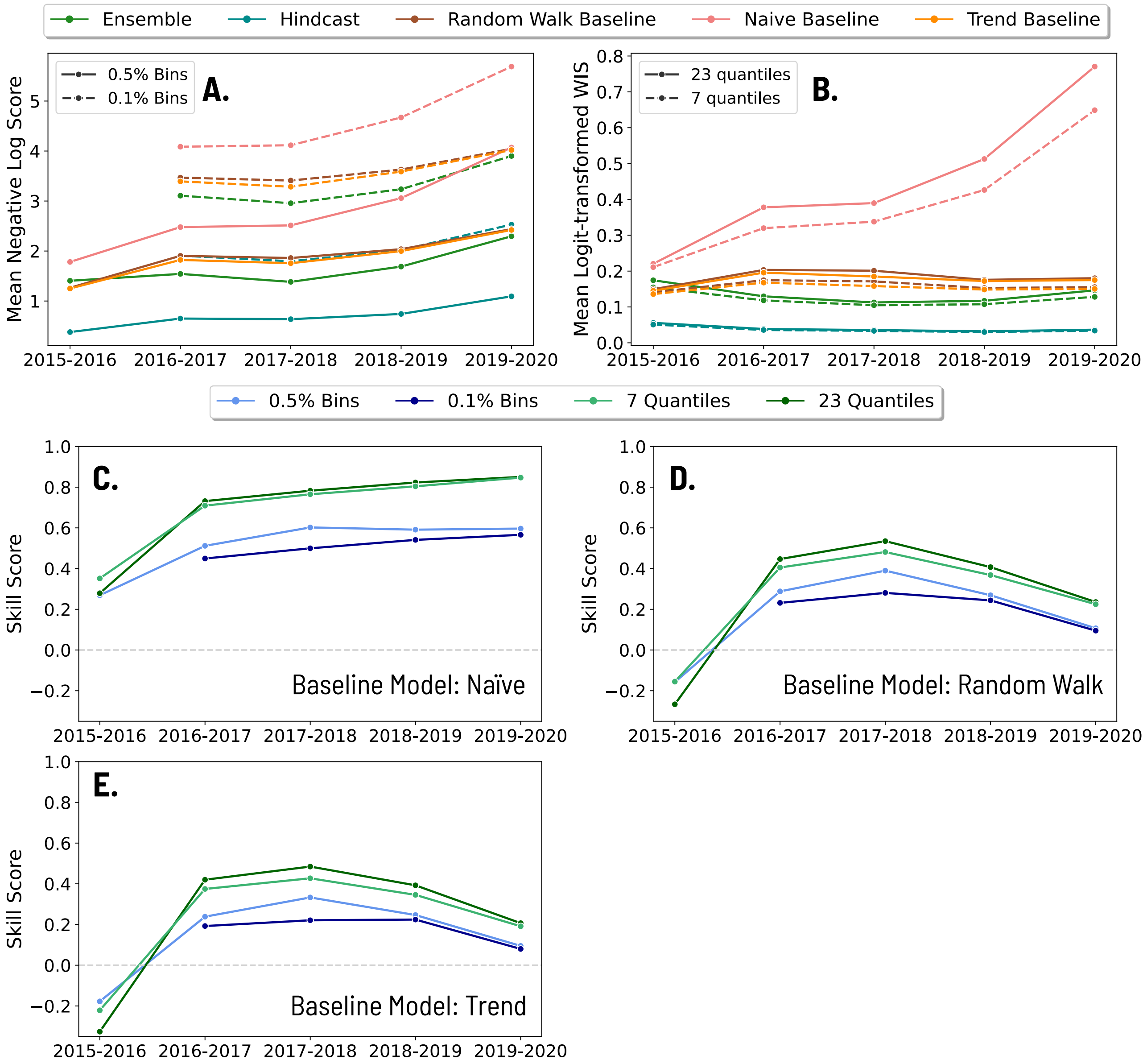


Figure S1: **Comparison of ILI forecast performance for binned probability distribution and quantile submission formats.** (A) Mean negative log score for each season of ILI forecasts for all models, comparing the scores using bigger bins (solid lines) and smaller bins (dashed lines) in the binned probability distribution format. Note there is no data for 2015-16 in the smaller bin format since forecasts were reported in the bigger bin format that season. (B) Mean log-transformed WIS for each season of ILI forecasts for all models, comparing the scores using 23 quantiles (solid lines) and 7 quantiles (dashed lines) in the quantile format. (C-E) Skill score of the ILI forecasts for each season with the 2 bin formats and 2 quantile formats. The reference model is (C) the naïve baseline, (D) the random walk baseline, and (E) the trend baseline.

Unlike the other disease targets, influenza-like illness (ILI) forecasts were submitted as binned probability distributions, where each bin represents the probability of the observed data falling within a specified range of % outpatient visits due to ILI. Early seasons, i.e. 2015-16, reported forecasts in 0.5% bin widths, while later seasons adopted narrower 0.1% bins. Moreover, the COVID-19 case forecasts used only 7 quantiles to describe the predictions, rather than the 23 used by the other COVID-19 and influenza forecast challenges. To assess the sensitivity of format and scoring choices, we compared scores across multiple configurations: the log score of forecasts with different bin sizes (0.5% and 0.1%) and the weighted interval score (WIS) with different quantile sets (7 and 23 quantiles).

We focused on the ILI forecasts, converting them to each of these formats to compare the resulting scores and skill scores. ILI is used as the test case since it is the only target available in the binned format. To assess the bin width, we aggregated the narrower 0.1% bins to the wider 0.5% width by summing the probabilities of consecutive bins within each 0.5% interval for the ILI forecasts from the 2016-17 season onward.

To convert the binned forecasts to quantiles, we computed the cumulative probability across all bins, and performed linear interpolation within the bins to identify the ILI value corresponding to each specified quantile level. The binned forecasts used the 0.5 or 0.1% bins from 0 to 13%, followed by a single bin spanning 13 to 100% (in 2014-15, 1% bins from 0-10%, then 10-100% bin) outpatient ILI visits that absorbs all remaining probability mass. Instead of interpolating linearly across this wide bin, which can cause extraordinarily high ILI estimates in the upper quantiles, we calculated the local CDF slope from the last narrow bin and extrapolate this slope forward to find where the CDF reaches the remaining quantiles. This preserves the quantile behavior into the tail rather than uniformly stretching the probability mass across the giant bin, which could inflate extreme values.

We extracted the 7 quantile levels used by the COVID-19 case forecasts (2.5, 10, 25, 50, 75, 90, 97.5 %) and the 23 levels used by the other forecasting challenges (1, 2.5, 5, 10, 15,..., 85, 90, 95, 97.5, 99%).

Figs. S1 A & B illustrate the scores for the ensemble and the three benchmark models (hindcast, naïve, random walk, and trend baseline) with the negative log score (LS) for binned forecasts and the logit-transformed WIS for quantile forecasts, respectively. We found that the wider 0.5% bins produced overall lower (better) log scores across models relative to the narrower 0.1% bins. This is because wider bins aggregate more probability mass, so each model can place a higher probability in the bin containing the true outcome. For the quantile forecasts, the 7-quantile format led to better WIS values than the 23-quantile format. Because the 7 quantiles used are a subset of the 23, this primarily reflects that fewer quantiles reduce the number of penalties contributing to the WIS.

Using these scores, we computed the skill score for each forecast format and both reference models (Figs. S1C - E). While the skill scores across data formats share similar trends, they differ in magnitude, limiting direct comparison between skill scores computed by different scoring rules. We found the highest skill scores for the 23-quantile WIS, with minimal differences relative to the 7-quantile WIS. Skill scores with the LS were lower overall and differed by bin size, with the wider 0.5% bins having higher skill. Due to these discrepancies, we performed all final analyses for the ILI forecasts using the 23-quantile format. Since the 7 and 23 quantile formats produce similar skill scores, COVID-19 case forecasts are included using the 7-quantile format, while all other disease outcomes are represented by 23 quantiles.

### 4 Additional Horizon-Specific Analyses

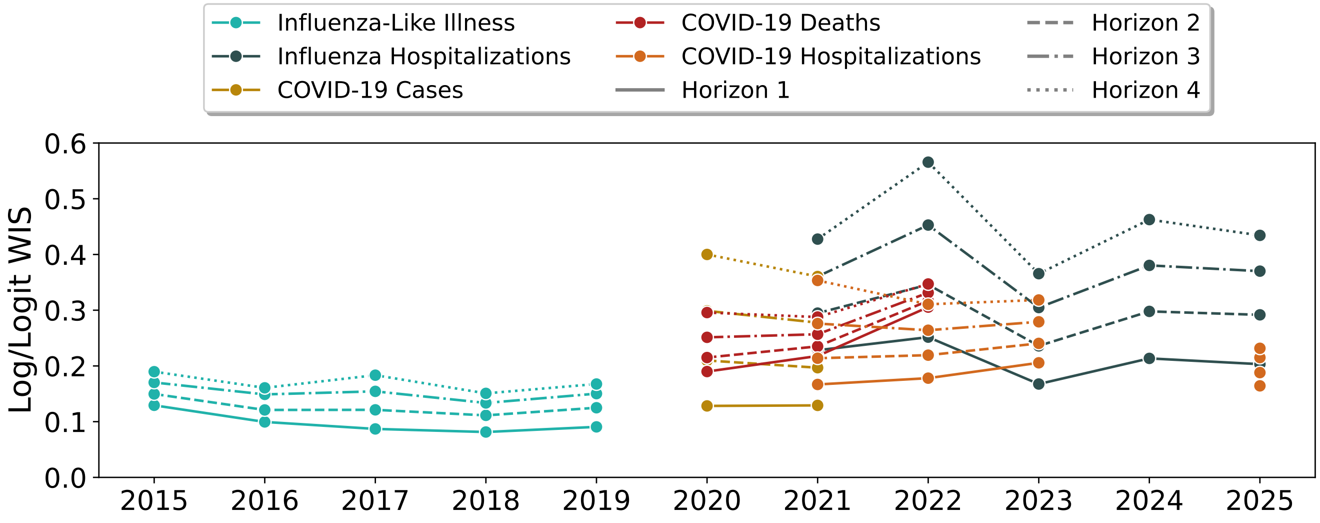


Figure S2: **Ensemble forecast weighted interval scores across seasons, disease outcomes, and forecast horizons.** Mean log-transformed WIS (logit for ILI) shown for the ensemble model across season for each disease outcome, with line style indicating the forecasting horizon.

Fig. S2 shows how the log/logit WIS of the ensemble changes over time for each horizon. For all disease outcomes, the first horizon (1) had the best (lowest) WIS values, and the last horizon (4) had the worst scores. This shows that performance degrades at longer horizon, corroborating past work on the impact of forecast horizon [4, 5].

Fig. S3 shows the ensemble skill score across horizons and disease outcomes relative to each of the three reference models. Relative to the naïve model, ensemble skill degrades at long horizons for all disease outcomes as in Fig. S2. The behavior was more varied for the RW and trend baselines. For some challenges the skill score decreased at longer horizons (e.g., COVID-19 hospitalizations and cases) while for others, skill increased at longer horizons (e.g., ILI). The behavior relative to the RW and trend baselines is a result of the interplay between how the ensemble and baseline scores both degrade at long horizons. Since the naïve model had stable performance across horizons (Fig. 2), the declining ensemble skill reflects higher ensemble WIS at longer horizons (Fig. S2).

### 5 Impact of the Hindcast on Forecast Skill

In previous evaluation of infectious disease forecasts (relative WIS [6, 7]) and other fields, the skill score has been calculated assuming no uncertainty in the data, assigning a perfect score, $S_{optimal} = 0$ [8, 9]. In Fig. S4, we compared how the interpretation of forecast skill changes with the inclusion of the hindcast, which accounts for variation in the observed data. While the two skill scores had similar behavior, the incorporation of the hindcast consistently increases the skill score across seasons and disease outcomes. This is a result of the hindcast changing the upper bound of forecast

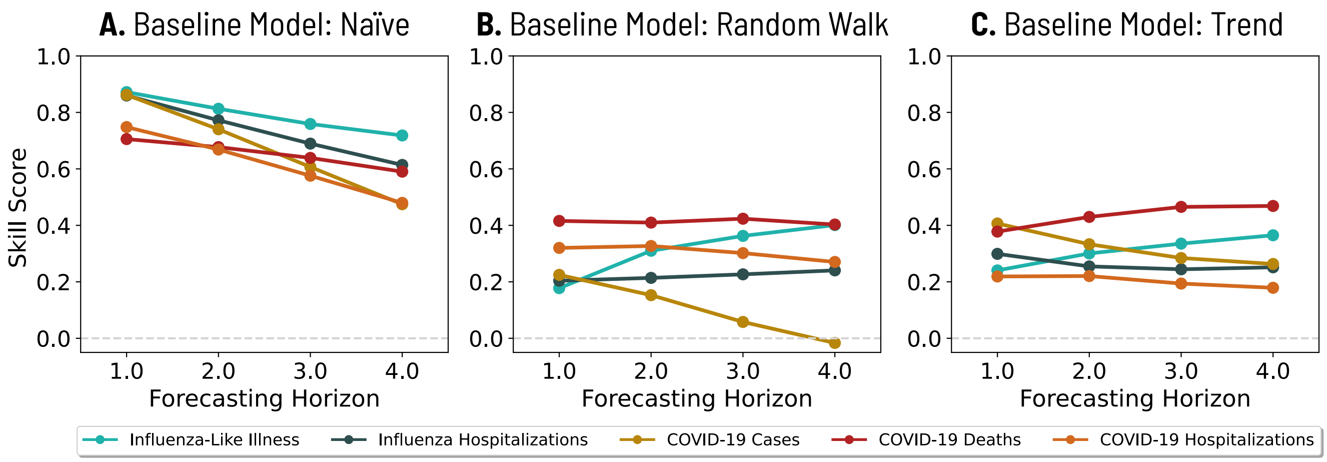


Figure S3: **Ensemble skill score at each horizon and disease outcome aggregated across locations and seasons.** The skill score was computed relative to the (A) naïve, (B) random walk, and (C) trend forecasts.

performance, where a forecast similar to the hindcast is more achievable than a forecast with a WIS of 0.

### 6 Submitted Baseline Comparison

We selected reference models which differ from the original baseline models produced for each forecast challenge. The ILI forecast baseline used seasonal patterns of historical data to create a forecast based on ILI activity in the same week in past years [10]. All COVID-19 and influenza hospitalization forecasts used the same baseline, which forecasts forward the most recently observed data point, with expanding uncertainty for later horizons based on historical count differences for consecutive weeks (a non-log-transformed version of our random walk baseline) [11].

We benchmarked forecast skill scores for the hub-generated ensemble and baseline forecasts in Fig. S5. As shown elsewhere, the ensemble model always outperformed the submitted baseline model. Relative to the reference models used here, the hub baseline models performed better than the naïve model, with the exception of the COVID-19 daily hospitalization predictions. Better relative performance is a reflection of the hub baseline models weighting recent data. The poor performance by the COVID-19 daily hospitalizations likely represents the increased noise in the surveillance timeseries resulting in increased uncertainty for the hub baseline model relative to other disease targets. The random walk (RW) model outperformed the original challenge baseline for all seasons and outcomes, likely due to differences in uncertainty from the log transformation. Similarly, the trend baseline performed better than the submitted baseline across seasons and outcomes, with the exception of COVID-19 cases and deaths in 2020, where performance was nearly equivalent. This demonstrates how the RW and trend baselines are a stricter benchmark for forecast performance than other commonly used reference models.

The hub baselines, like the RW and trend baselines, present a challenge for analyzing skill by horizon. The pairwise relative WIS is a version of the skill score that has been used to evaluate

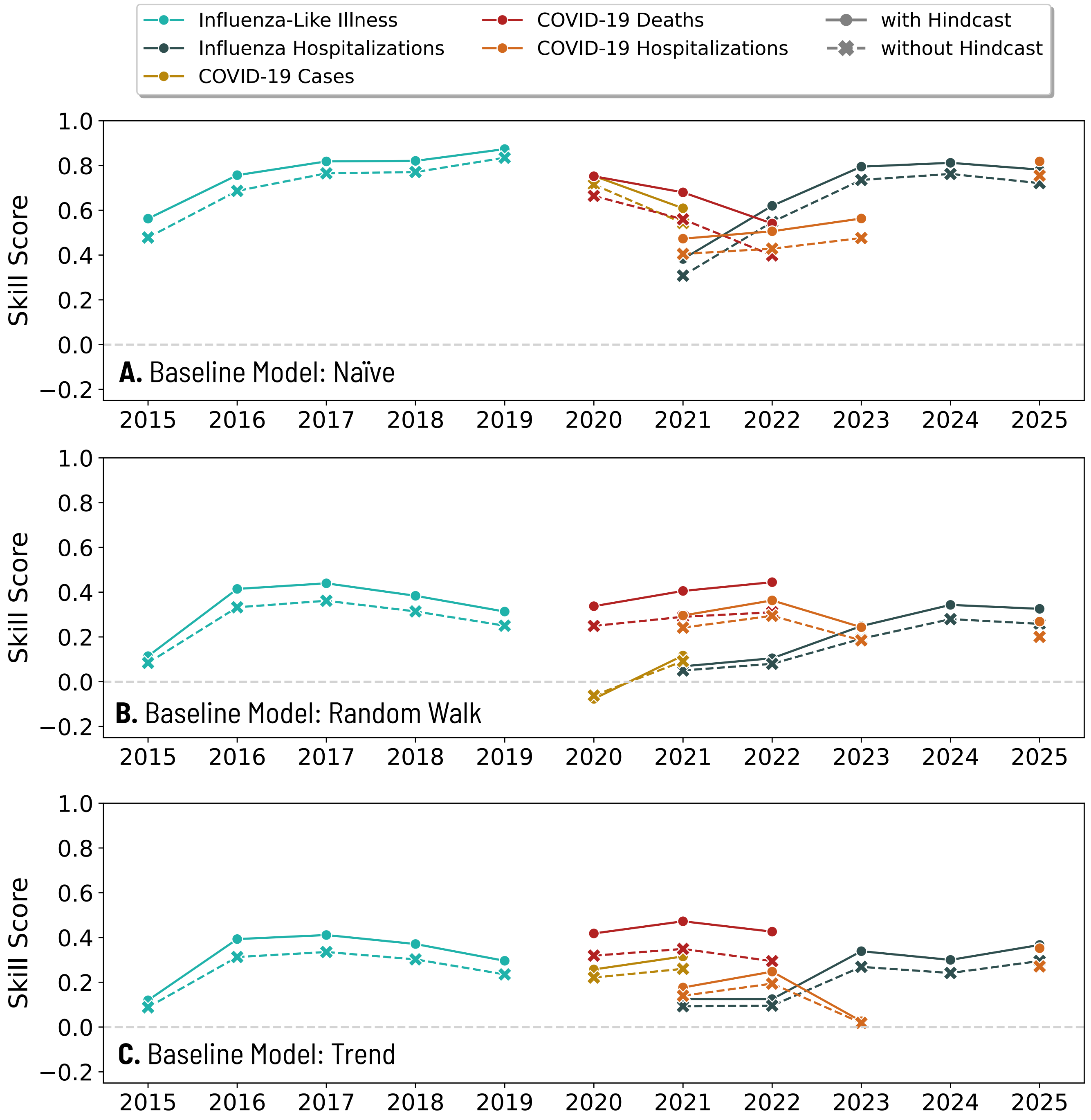


Figure S4: **Comparison of ensemble skill scores with perfect outcomes versus hindcasts as the optimal forecast.** Skill score for the ensemble model across season relative to each reference model (A) naïve baseline, (B) random walk baseline, and (C) trend baseline, comparing how the hindcast influences the skill score. The dashed line shows the skill score where the optimal score is assumed to be perfect, i.e., 0. The solid line shows the skill score with the hindcast.

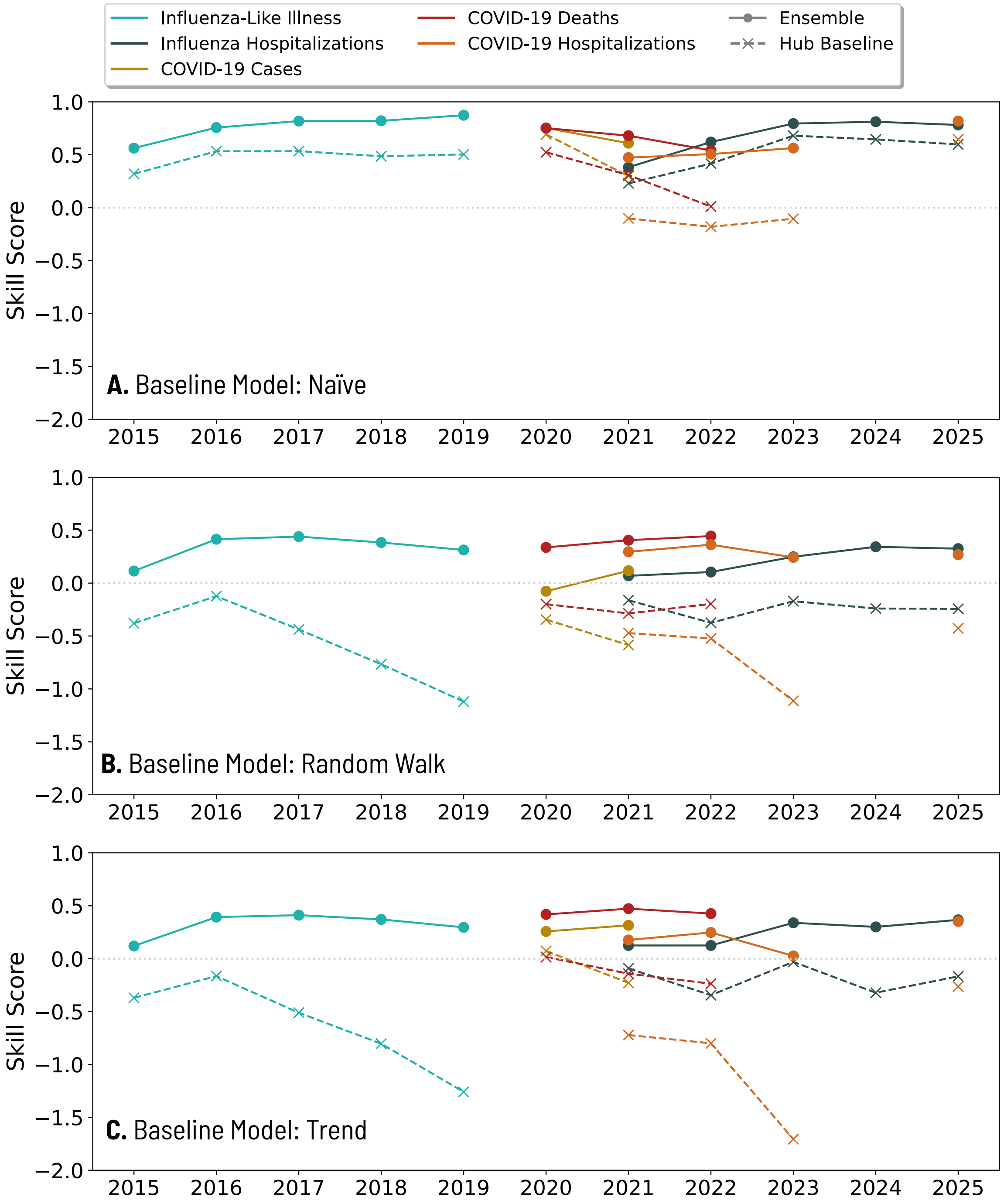


Figure S5: **Assessment of hub-generated baseline skill score across seasons and forecasting targets relative to each of the three baseline models developed here.** Skill score for the hub-generated ensemble and hub-generated baseline forecasts across disease outcomes and seasons. Skill scores were computed relative to the three reference models: (A) naïve baseline, (B) the random walk baseline, and (C) the trend baseline. A skill above 0 indicates better performance by the ensemble or baseline than the corresponding reference model.

forecasts relative to the hub baselines [6, 7]. It is computed by taking the geometric mean of the pairwise WIS across all models and dividing by the same value for the baseline model. The final step of this calculation, scaling to the baseline, is equivalent to the skill score where the skill score $SS = 1 - \text{WIS}_f/\text{WIS}_{baseline}$, if the optimal model is assumed to be perfect, with a score of $S_{optimal} = 0$. Figure S6 shows the simplified relative WIS of the ensemble compared to the hub-generated baseline across outcome, season, and horizon ($\text{WIS}_{ensemble}/\text{WIS}_{baseline}$). This shows agreement with features we see with the skill score. Namely, the ensembles consistently performed better than the baseline model, and ILI and flu hospitalization forecasts improved over time. In addition, we find the counterintuitive behavior by horizon identified in Fig. 3. Here, influenza hospitalization and COVID-19 death forecasts had better relative WIS at long horizons. This follows from the faster degradation of the hub baseline model at long horizons compared to the ensemble, the same pattern seen with the RW and trend baselines.

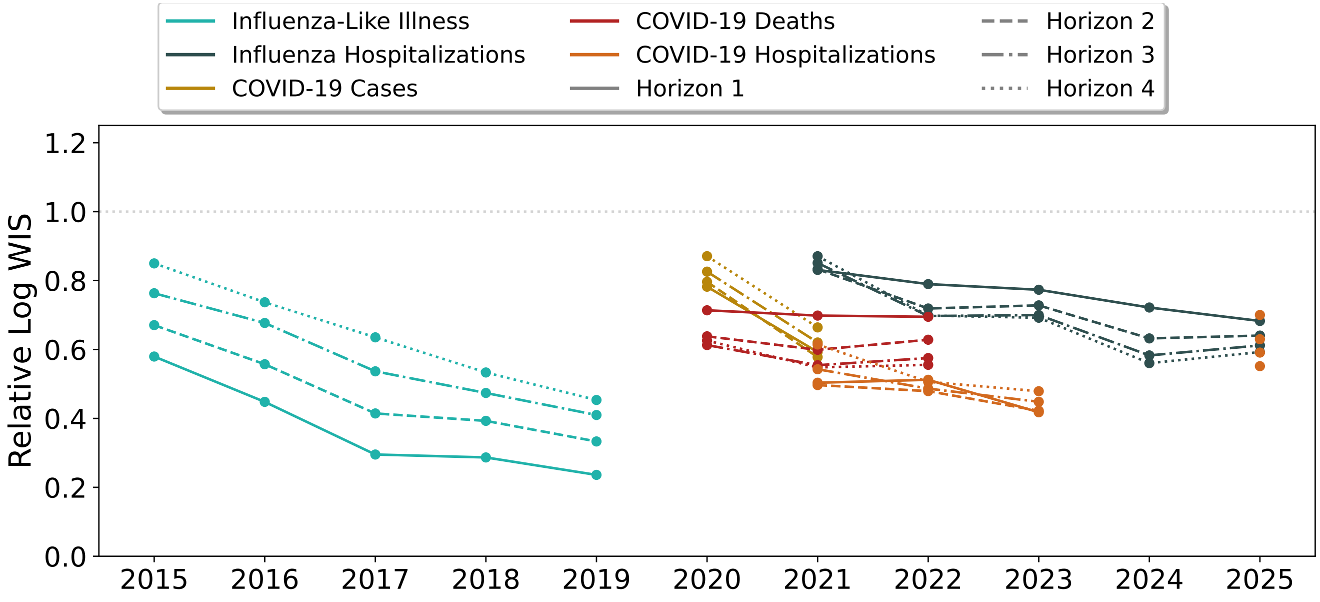


Figure S6: **Ensemble relative WIS compared to the hub-generated baseline models.** Relative WIS of the hub-generated ensemble against the hub-generated baseline model across seasons, forecasting horizons, and disease outcomes. A relative WIS below 1 represents better performance by the ensemble. The line style represents different forecasting horizons.

### 7 Baseline Model Selection

While we considered three reference models throughout this work (the naïve, random walk, and trend models), there are many different variations and methodologies of baseline models that have been used or proposed in other work [12, 13]. In addition to the existing hub baselines, we compared the performance of our baseline models with nine other baseline forecasts using the mean log-transformed WIS for each outcome and season (Fig. S7). The additional nine baseline models were proposed in [12] and included constant, marginal, kernel density (KDE), last similar date (LSD, seasonal baseline), ordinary least squares (OLS), increase-decrease-stable (IDS, variant of OLS with linear trend), ARMA, exponential smoothing (ETS), and season and trend decomposition with LOESS (STL) models. We considered the log-transformed version of these 9 models because we find that log-transformed baselines typically had improved performance compared to the corresponding non-transformed versions. We analyzed the forecasts directly generated by [12] for the seasons and outcomes shown here.

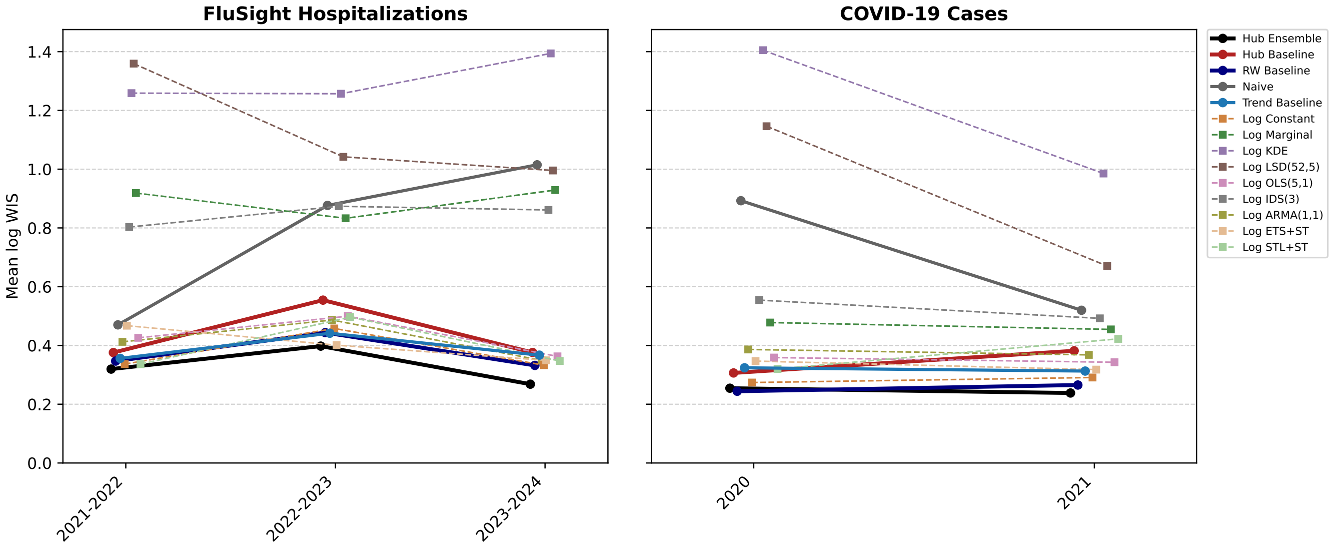


Figure S7: **Comparison of forecast performance for 14 different baseline models for influenza hospitalization and COVID-19 case forecasts.** Mean log-transformed WIS across seasons for influenza hospitalization and COVID-19 case forecasts for the ensemble and three baseline models considered in this work (highlighted), along with 10 additional baseline models proposed by others.

As in Fig. S5, the RW and trend models generally outperformed the hub baseline models. In addition, the RW baseline models had improved performance compared to most of the additional baseline models and the trend baseline was also consistently good. While forecast skill can be computed relative to any choice of reference model, this shows that the RW baseline, in particular, is relatively strict benchmark for forecast performance. While the naïve model had higher scores than the RW and trend baselines, it performed better than the Log KDE and Log LSD models. This demonstrates the benefit of the naïve approach over using a seasonal baseline and highlights the utility of simplistic reference models in assessing forecast skill.

### 8 Team Forecasts

While much of the analysis presented here focuses on the performance of the ensemble model of each forecasting hub, we also examined participation and the skill of individual models (Fig. S8). We found that for both influenza targets, the number of contributing models increased over time. The increase in teams was accompanied by an increase in the proportion of models with a positive skill (better than the reference model) relative to each of the three baseline models (naïve, RW and trend). For COVID-19 cases and deaths, patterns were mixed, with declining performance

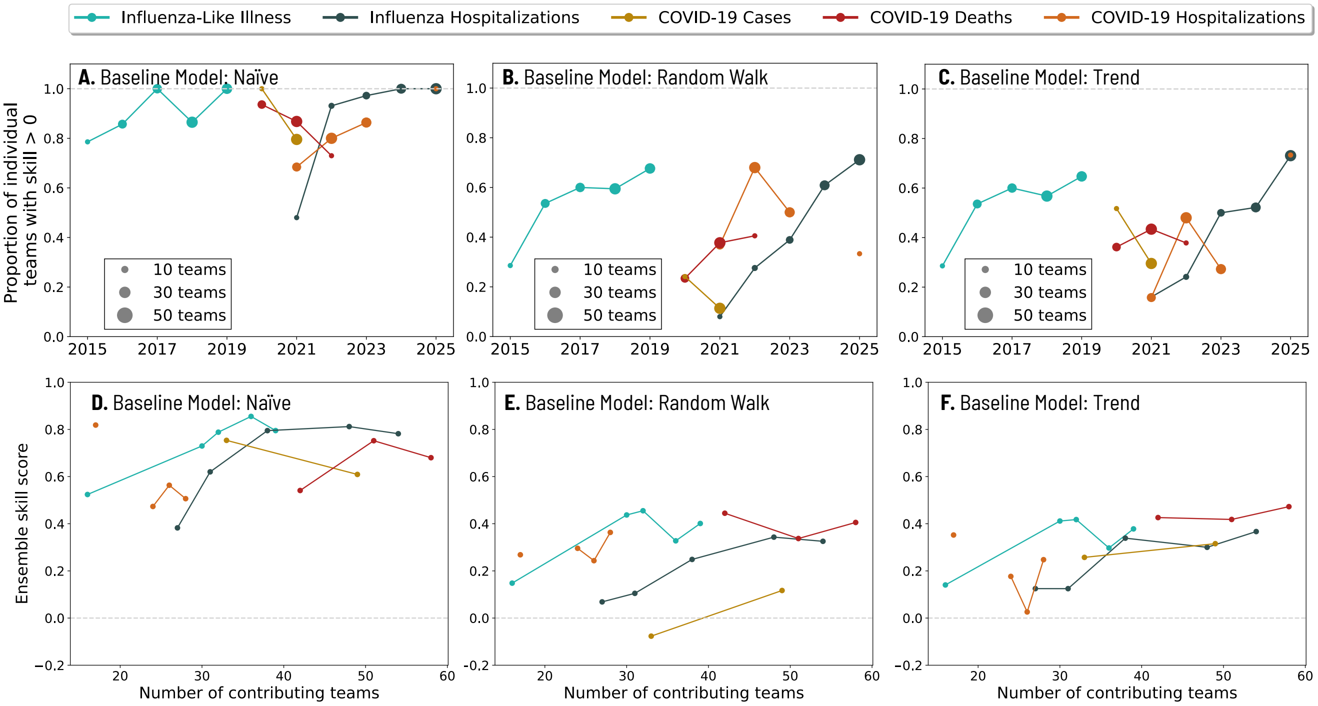


Figure S8: **Relative skill scores for individual modeling teams across seasons and correlation between participation and the ensemble skill score.** The proportion of teams per season and disease outcome with a positive skill score relative to the (A) naïve, (B) random walk, and (C) trend baseline models. Markers are sized by the number of contributing modeling teams. Ensemble skill score versus number of contributing modeling teams relative to the (D) naïve, (E) random walk, and (F) trend baseline models.

relative to the naïve forecasts, which follows the ensemble skill score. These patterns largely follow the behavior of the skill score for the ensemble, highlighting the importance of forecast quality at the individual model level. Notably, for the two ongoing forecasting challenges (COVID-19 and influenza hospitalizations), the most recent seasons each saw 100% of models achieving a skill score better than the naïve baseline model.

Comparing the relationship between the number of contributing teams and the ensemble skill score, we found that for both influenza targets, a higher number of models was generally correlated with better ensemble skill scores. For COVID-19 the results were less clear, likely reflecting higher variability in the data and in participation.

### References

[1] N. G. Reich, et al., Collaborative hubs: Making the most of predictive epidemic modeling, Am. J. Public Health 112 (6) (2022) 839–842. doi:10.2105/AJPH.2022.306831.
URL https://doi.org/10.2105/AJPH.2022.306831

[2] J. Bracher, On the multibin logarithmic score used in the flusight competitions, Proceedings of the National Academy of Sciences 116 (42) (2019) 20809–20810. doi:10.1073/pnas.1912147116.
URL https://www.pnas.org/doi/abs/10.1073/pnas.1912147116

[3] T. Gneiting, A. E. Raftery, Strictly proper scoring rules, prediction, and estimation, Journal of the American Statistical Association 102 (477) (2007) 359–378. doi:10.1198/016214506000001437.
URL https://doi.org/10.1198/016214506000001437

[4] C. S. Lutz, M. P. Huynh, M. Schroeder, S. Anyatonwu, F. S. Dahlgren, G. Danyluk, D. Fernandez, S. K. Greene, N. Kipshidze, L. Liu, et al., Applying infectious disease forecasting to public health: a path forward using influenza forecasting examples, BMC Public Health 19 (1) (2019) 1–12.

[5] C. Viboud, A. Vespignani, The future of influenza forecasts, Proceedings of the National Academy of Sciences 116 (8) (2019) 2802–2804. doi:10.1073/pnas.1822167116.
URL https://www.pnas.org/doi/abs/10.1073/pnas.1822167116

[6] E. Y. Cramer, et al., Evaluation of individual and ensemble probabilistic forecasts of covid-19 mortality in the united states, Proc. Natl. Acad. Sci. U.S.A. 119 (15) (2022) e2113561119. doi:10.1073/pnas.2113561119.

[7] E. Howerton, L. Contamin, et al., Evaluation of the US COVID-19 scenario modeling hub for informing pandemic response under uncertainty, Nature Communications 14 (1) (2023) 7260. doi:10.1038/s41467-023-42680-x.
URL https://doi.org/10.1038/s41467-023-42680-x

[8] A. H. Murphy, Skill Scores Based on the Mean Square Error and Their Relationships to the Correlation Coefficient (Dec. 1988). doi:10.1175/1520-0493(1988)116<2417:SSBOTM>2.0.CO;2.
URL https://journals.ametsoc.org/view/journals/mwre/116/12/1520-0493_1988_116_2417_ssbotm_2_0_co_2.xml

[9] E. Wheatcroft, Interpreting the skill score form of forecast performance metrics, International Journal of Forecasting 35 (2) (2019) 573–579. doi:10.1016/j.ijforecast.2018.11.010.
URL https://www.sciencedirect.com/science/article/pii/S0169207019300093

[10] C. J. McGowan, M. Biggerstaff, M. Johansson, K. M. Apfeldorf, M. Ben-Nun, L. Brooks, M. Convertino, M. Erraguntla, D. C. Farrow, J. Freeze, S. Ghosh, S. Hyun, S. Kandula, J. Lega, Y. Liu, N. Michaud, H. Morita, J. Niemi, N. Ramakrishnan, E. L. Ray, N. G. Reich, P. Riley, J. Shaman, R. Tibshirani, A. Vespignani, Q. Zhang, C. Reed, Collaborative efforts

to forecast seasonal influenza in the United States, 2015–2016, Scientific Reports 9 (1) (2019) 683. doi:10.1038/s41598-018-36361-9.
URL https://www.nature.com/articles/s41598-018-36361-9

[11] E. L. Ray, L. C. Brooks, J. Bien, M. Biggerstaff, N. I. Bosse, J. Bracher, E. Y. Cramer, S. Funk, A. Gerding, M. A. Johansson, A. Rumack, Y. Wang, M. Zorn, R. J. Tibshirani, N. G. Reich, Comparing trained and untrained probabilistic ensemble forecasts of covid-19 cases and deaths in the united states, International Journal of Forecasting 39 (3) (2023) 1366–1383. doi:https://doi.org/10.1016/j.ijforecast.2022.06.005.
URL https://www.sciencedirect.com/science/article/pii/S0169207022000966

[12] M. Stapper, S. Funk, Mind the baseline: The hidden impact of reference model selection on forecast assessment, medRxiv (2025). arXiv:https://www.medrxiv.org/content/early/2025/09/03/2025.08.01.25332807.full.pdf, doi:10.1101/2025.08.01.25332807.
URL https://www.medrxiv.org/content/early/2025/09/03/2025.08.01.25332807

[13] E. Suez, S. J. Fox, Basic baseline model design choices can substantially influence performance in collaborative forecast hubs, medRxiv (2026). doi:10.64898/2026.03.18.26348748.
URL https://www.medrxiv.org/content/early/2026/03/20/2026.03.18.26348748
